# COVID-19 Impact on Individuals with Type 1 and Type 2 Diabetes: Comparison of Pre- and Post-COVID-19 Hospitalizations for Diabetes-Related Amputation

**DOI:** 10.64898/2026.01.02.25342941

**Authors:** Yelena Ionova, Lixian Zhong, Ruben Vargas, Yanlei Ma, Leslie Wilson

## Abstract

**Background:** The COVID-19 pandemic disrupted healthcare services, potentially affecting diabetes management and complications.

**Objective:** To investigate the impact of the pandemic on lower extremity amputation (LEA) rates among individuals with type 1 and type 2 diabetes mellitus, focusing on social determinants of health.

**Methods:** A retrospective observational cohort study using de-identified claims data from a large U.S. health plan. LEA rates were compared before and after the onset of the COVID-19 pandemic using interrupted time series analysis.

**Results:** Individuals with type 2 diabetes experienced an initial decline in LEA rates followed by a significant increase (p=0.022) as delayed care needs were addressed. Individuals with type 1 diabetes showed no significant fluctuations in amputation rates. Social determinants were significantly associated with changes in LEA rates among individuals with type 2 diabetes. Lower-income (≤$40,000/year) and less educated individuals experienced significant increases in amputation rates (p=0.027 and p=0.043, respectively). Individuals aged 45-64 years showed a significant increase in LEA rates (p=0.013), while those aged 18-44 experienced a decrease (p=0.017). Metropolitan residents saw significant increases in LEA rates (p=0.021).

**Conclusions:** The COVID-19 pandemic significantly disrupted healthcare access for individuals with type 2 diabetes, leading to increased LEA rates. Social determinants of health exacerbated existing disparities in diabetes outcomes. These findings underscore the need for targeted interventions to address healthcare disparities, especially during public health crises.

## INTRODUCTION

Diabetes is a highly prevalent chronic condition affecting approximately 11.6% of the U.S. population. It imposes significant economic costs and is associated with severe morbidity and mortality.^1,2^ Effective diabetes management is burdensome, requiring regular clinical care, adherence to medication regimen, and diligent lifestyle monitoring. Without adequate healthcare access and adherence to rigorous diabetes management plan, individuals with diabetes are at risk of developing diabetes-related complications. Among these, poorly controlled diabetes can lead to lower limb complications such as diabetic foot ulcers, poor circulation, and subsequent amputations.^3^

The emergence of Coronavirus disease 2019 (COVID-19) as a global pandemic impeded access to healthcare as clinics transitioned to virtual care, hospitals enforced capacity, and fear of COVID-19 infection reduced the number of patients seeking care. The shift in healthcare access was compounded by the economic challenges brought about by the pandemic. Increasingly people were unable to afford insulin and other medications as unemployment increased with COVID-19. American Diabetes Association estimates that 15% of individuals with diabetes who use pumps and continuous glucose monitors delayed refilling prescriptions largely due to financial strain, and 12% lost their previous insurance within the first nine months of the pandemic.^4^

The impact of the pandemic on healthcare access was not uniform across population groups. Research suggests that minority groups have historically faced disparities in healthcare access, and these disparities were likely exaggerated during the early years of the COVID-19 pandemic.^5^ For individuals with chronic diseases like diabetes, delays in care can lead to more severe outcomes.

Our objective is to examine the overall changes in access to hospital care and the resulting changes in outcomes during the first year of the COVID-19 pandemic. Additionally, we will determine any disproportionate effects on minority groups and outcomes in individuals with diabetes across pre- and post-COVID-19 periods. Specifically, we aim to assess whether the changes in access to care affected the rates of lower extremity amputations in persons with diabetes and how these outcomes differed by equity groups one-year prior and one-year following the onset of COVID19.

## RESEARCH DESIGN AND METHODS

### Design and Data Source

This study employs a retrospective observational cohort design, utilizing both pre-post and interrupted time series (ITS) methodologies. We utilized de-identified claims data from commercial and Medicare Advantage enrollees, sourced from the OptumLabs Data Warehouse (OLDW) National and PanTher (eHR-derived data) View Files. The OLDW is a comprehensive database that integrates administrative claims and health records for over 160 million de-identified individuals dating back to 1993 and is provided by a large and geographically diverse U.S. health plan.

### Study Population

Two distinct samples, comprising individuals with diabetes mellitus type 1 (T1DM) and type 2 (T2DM), were identified from January 1^st^, 2016, to March 1^st^, 2019. The inclusion criteria required at least two claims with ICD-10 codes E10 (T1DM) or E11 (T2DM), or a single diagnosis claim accompanied by a record of diabetes-related medication prescription. More detailed descriptions of the sample selection process and the inclusion/exclusion criteria are documented in previous publications.^6,7^ The final sample consisted of 103,290 individuals with T1DM and 711,951 individuals with T2DM (Supplemental Figure 1).

### Variables

#### Exposure

The onset of the COVID-19 pandemic serves as the intervention or exposure of interest for this study. March 2020 was selected as the commencement of the pandemic, aligned with the World Health Organization’s (WHO) declaration of COVID-19 as a global pandemic.^8^

#### Covariates

The analysis examined various demographic and equity variables to identify disparities in healthcare access and outcomes. The covariates included age, categorized into three groups (18-44, 45-64, and 65+ years); sex (male and female); race/ethnicity (White, Black, Hispanic, Asian, and Other); socioeconomic status, categorized into five ranges based on income levels (<$40,000, $40,000-$74,999, $75,000-$124,999, $125,000-$199,999, and >$200,000); education level (less than high school, high school, some college, and bachelor’s degree or higher); and geographic location (metropolitan, micropolitan, small town, and rural).

#### Healthcare Access

Healthcare access was assessed by measuring changes in the monthly hospitalization rate per person, comparing the year prior to the onset of COVID-19 pandemic (February 2019 to February 2020) to the first year after the pandemic began (March 2020 to February 2021).^6,7^

#### Outcomes

The primary outcome of interest was the rate of lower extremity amputations (LEA), determined using procedure codes for LEA in patients hospitalized with diabetes that were defined by criteria established by the Agency for Healthcare Research and Quality (AHRQ).^9^ Patients admitted with any diagnosis of diabetes and any lower-extremity amputation procedure (except toe amputations) were included in the analysis. To avoid confounding, hospital admission with confirmed and probable/suspected COVID-19 diagnoses were excluded. The rate of amputation was calculated monthly as the ratio of the number of hospital admissions with amputations to the total number of enrolled patients with type 1 and type 2 diabetes, respectively.

### Statistical Analysis

Descriptive statistics summarized baseline demographic characteristics of the study population. Interrupted time series (ITS) analysis was employed to assess significant differences in amputation outcomes across two distinct time periods, controlling for temporal effects. The ITS regression model, applied to the monthly amputation rate, incorporated terms for time trend, COVID-19 onset intervention, and their interaction. Regression coefficients were used to estimate the intervention’s effect, adjusting for the underlying time trend. Data extraction from the OptumLabs database utilized Structured Query Language (SQL), and analyses were conducted using R version 4.3.2. Approval for this study was granted by the University of California, San Francisco Institutional Review Board (IRB).

## RESULTS

### Demographics and Social Characteristics

The demographics have been described in previous studies,^6,7^ but in general for both diabetes types, consist of majority whites with some college education, living in a metropolitan setting (Supplemental Table 1). Annual income levels are more representative across a range from less than $40,000 to $125,000.

### Health Care Access

Changes in hospitalization rates for both type 1 and type 2 diabetes with the onset of COVID-19 have been reported elsewhere.^6,7^ In general, we found that for both types of diabetes, there was an immediate and significant drop in hospitalization rate per 100,000 persons during the first month following the onset of COVID-19 pandemic. For type 1 diabetes, this initial drop was followed by a significant rise in hospitalization rates. This increase continued until the hospitalization rate per 100,000 persons reached the level of the calculated expected rate, indicating a return to pre-pandemic levels as delayed care needs were addressed. For type 2 diabetes, the initial drop in hospitalization rates was also followed by a significant rise. However, the hospitalization rate for type 2 diabetes patients reached and surpassed the calculated expected rate by the end of the first year of the COVID-19 pandemic. This result demonstrated that there was a significant change in access to hospitalizations for those with diabetes and this rate increased significantly to account for the delayed care available at that time.

### Amputation Outcomes

Following the analysis of hospitalization rates that are described in detail in previous publications,^6,7^ we examined whether the rate of lower extremity amputations (LEA) changed in association with the delayed care observed during the first year of the pandemic.

For those with type 1 diabetes, there was a sharp drop in amputations during the first month of the COVID pandemic start, but this drop was not significant. The rise in amputation rate during the first post-COVID year was also not significant, and followed the same slope as that predicted, but never reaching the pre-pandemic amputation levels, suggesting that the rate of amputations for type 1 diabetes patients remained relatively stable throughout the study period (Figure 1a).

**Figure 1:**
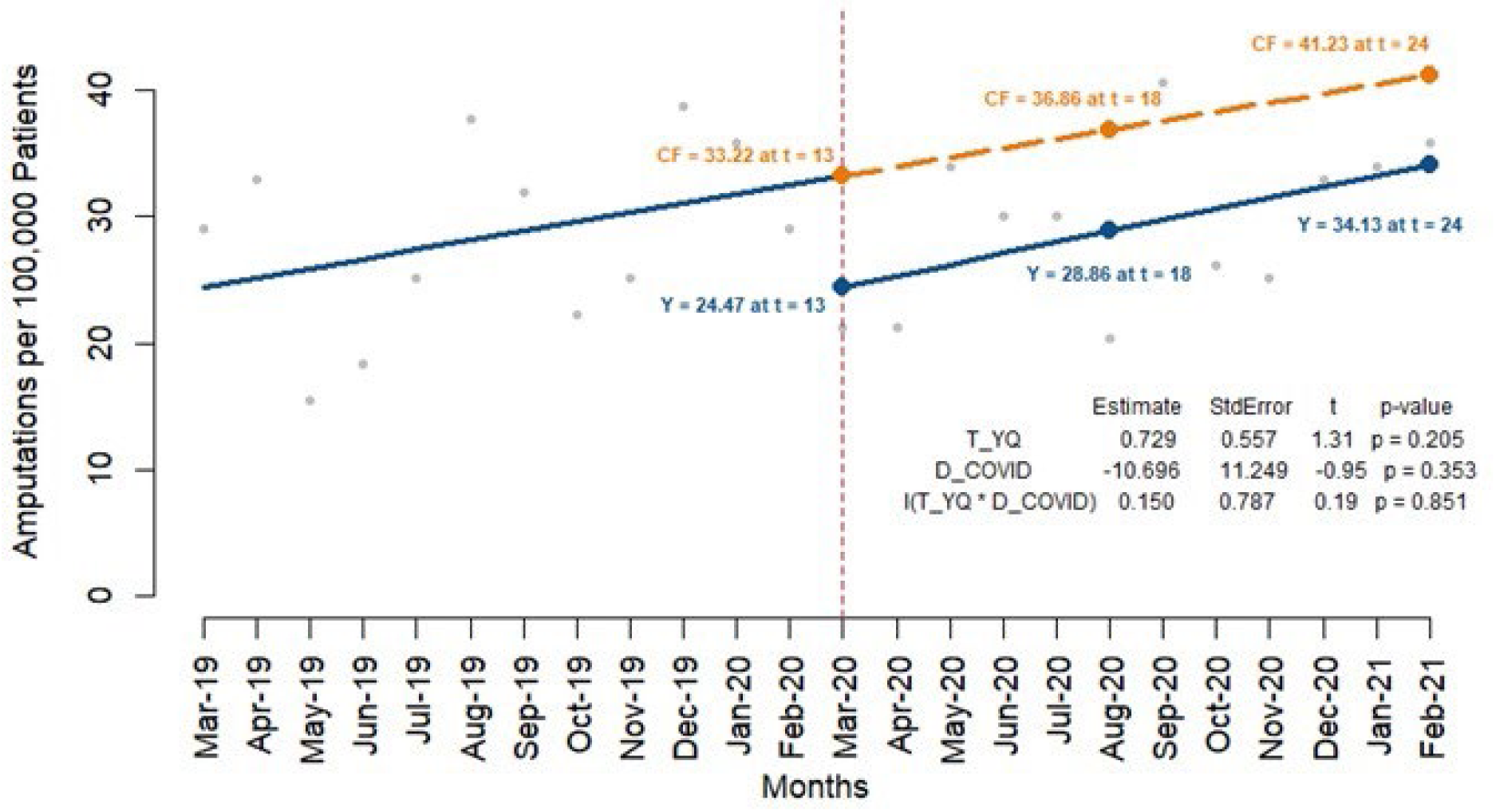
Lower Extremity Amputation Rate per 100,000 patients during first year pre- and post-COVID-19 Pandemic. **Figure 1a:** Lower Extremity Amputation Rate per 100,000 Type 1 Diabetes patients during first year pre- and post-COVID-19 Pandemic Relative drop = 24.7%

**Figure 1b:**
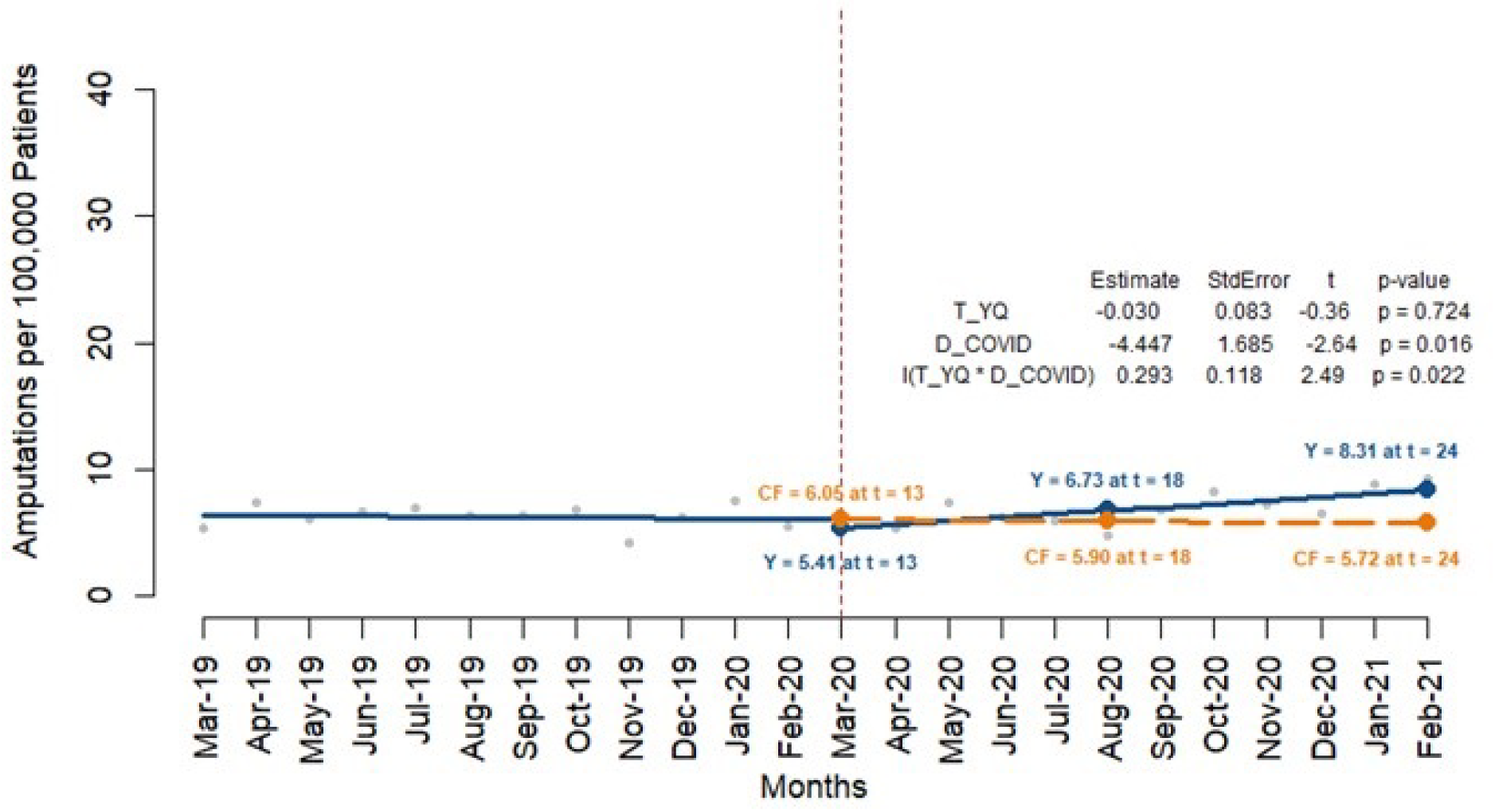
Lower Extremity Amputation Rate per 100,000 Type 2 Diabetes patients during first year pre- and post-COVID-19 Pandemic Relative drop = 10.9%

In contrast, the analysis for type 2 diabetes patients indicated significant changes in amputation rates per 100,000 patients due to the pandemic. There was an immediate absolute drop in amputations during the first month onset of the COVID pandemic (−4.447, p=0.016) with a relative drop of 45.1% amputations per 100,000. Over the first pandemic year there was a significant continued rise in monthly rate of hospitalized amputation (0.293, p = 0.022), resulting in a higher amputation rate than estimated by the end of the first pandemic year (actual = 8.31; predicted = 5.72) (Figure 1b). These findings highlight a significant disruption in healthcare access for type 2 diabetes patients with an initial decline in amputations followed by a compensatory increase as delayed care needs were addressed.

### Effects of Social Determinants on Amputation Outcomes

Interestingly, the analysis revealed fewer disparities among demographic and equity groups in type 1 diabetes compared to type 2 diabetes. When stratifying by demographic and equity factors such as age, race, education, income, and geographic location, there were no statistically significant differences in amputation rates among type 1 diabetes patients pre- and post-COVID-19 (Supplemental Figures 2-6).

In contrast, our analysis revealed significant differences in lower extremity amputation rates among patients with type 2 diabetes when stratified by various social determinants of health. For patients aged 18-44 years, there was a significant decrease in amputation rates post-pandemic (−1.000, p=0.017) (Figure 2a). In contrast, patients aged 45-64 years exhibited a significant increase in amputation rates (0.587, p=0.013) (Figure 2b), while no statistically significant changes were observed for patients aged 65 and over (Figure 2c).

**Figure 2:**
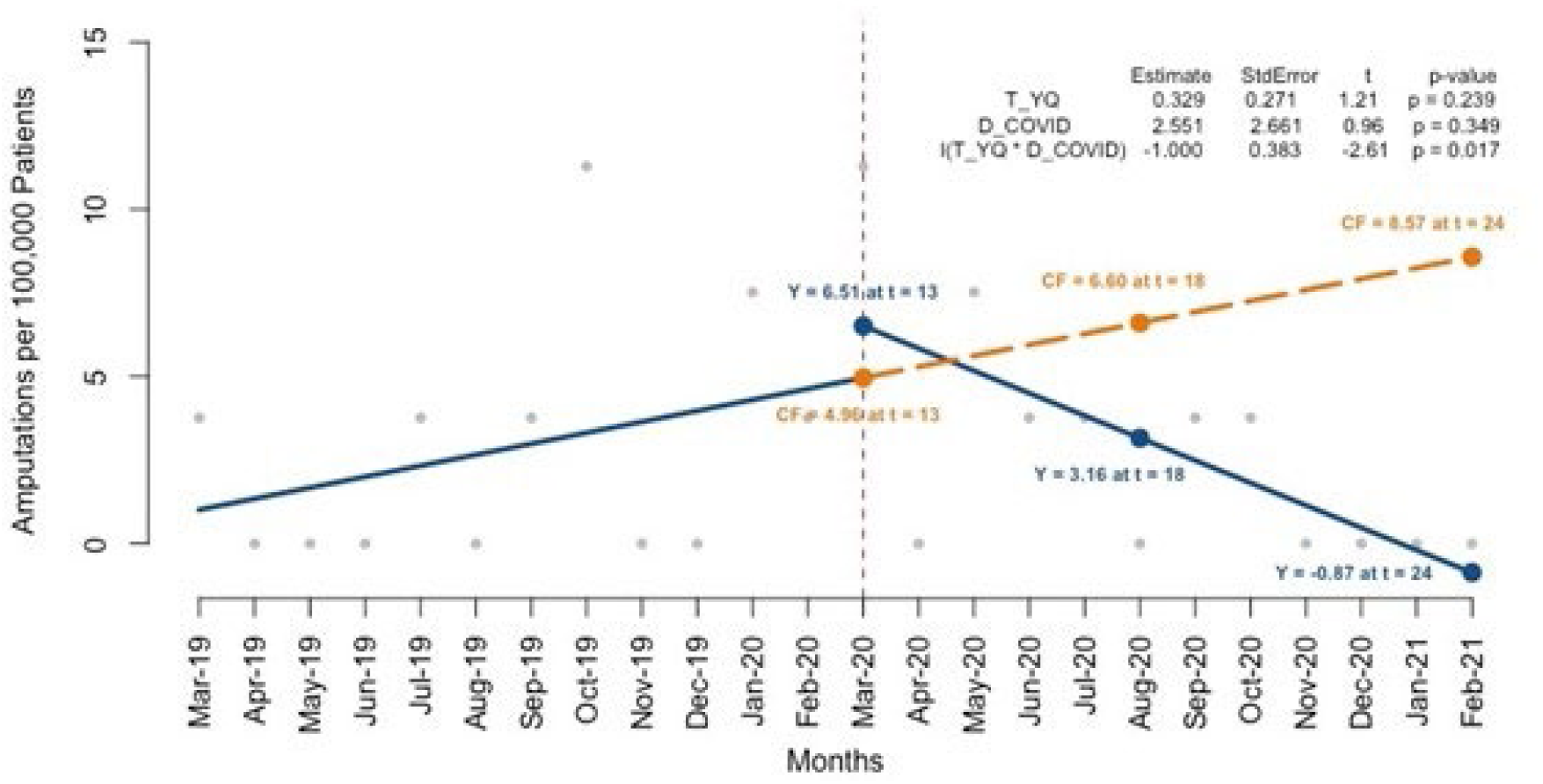
Amputations per 100,000 Type 2 Diabetes Patients by Age. **Figure 2a:** Amputations per 100,000 Type 2 Diabetes Patients Aged 18-44 Relative increase = 40.6%

**Figure 2b:**
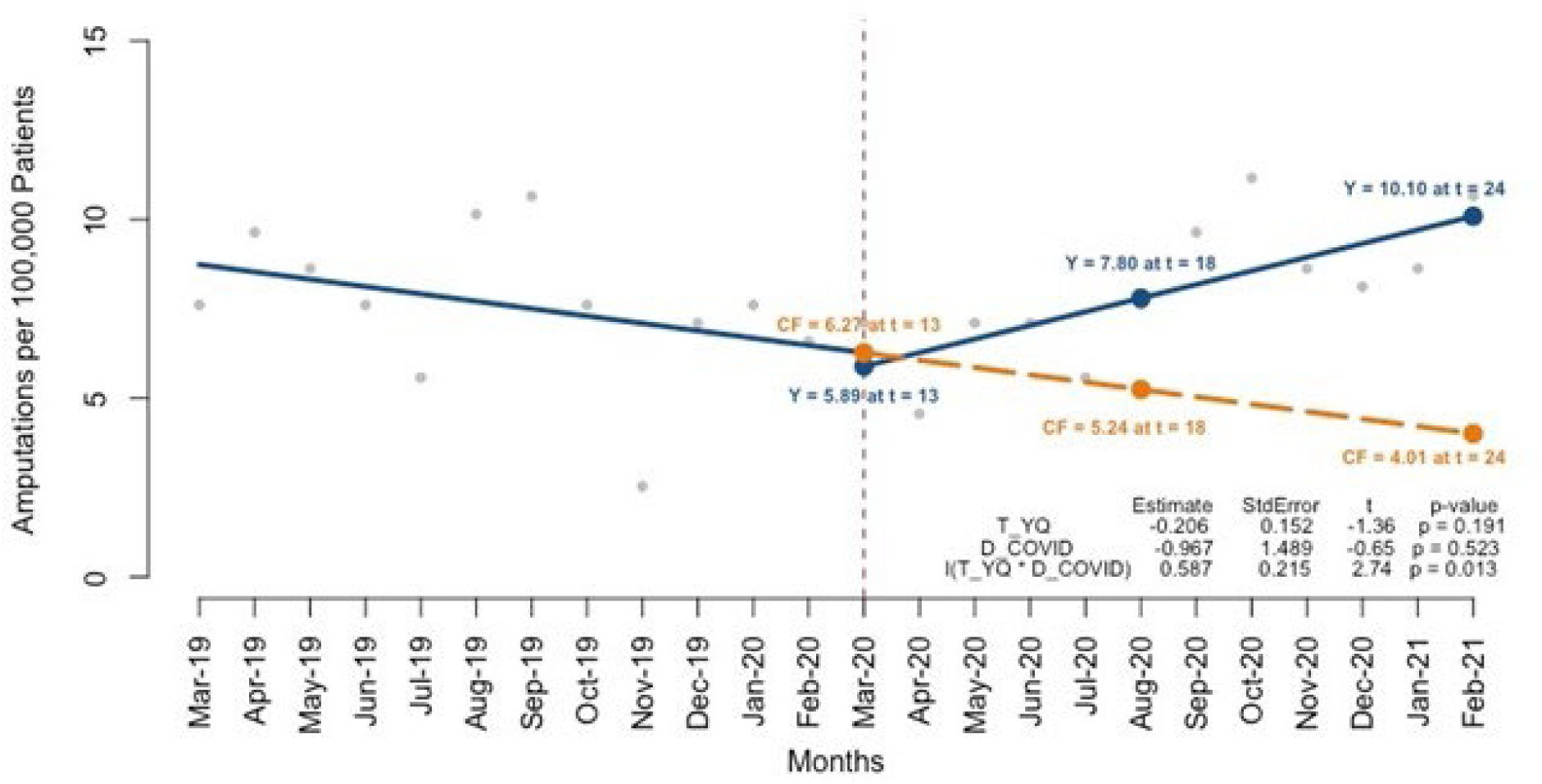
Amputations per 100,000 Type 2 Diabetes Patients Aged 45-64 Relative drop = 9.0%

**Figure 2c:**
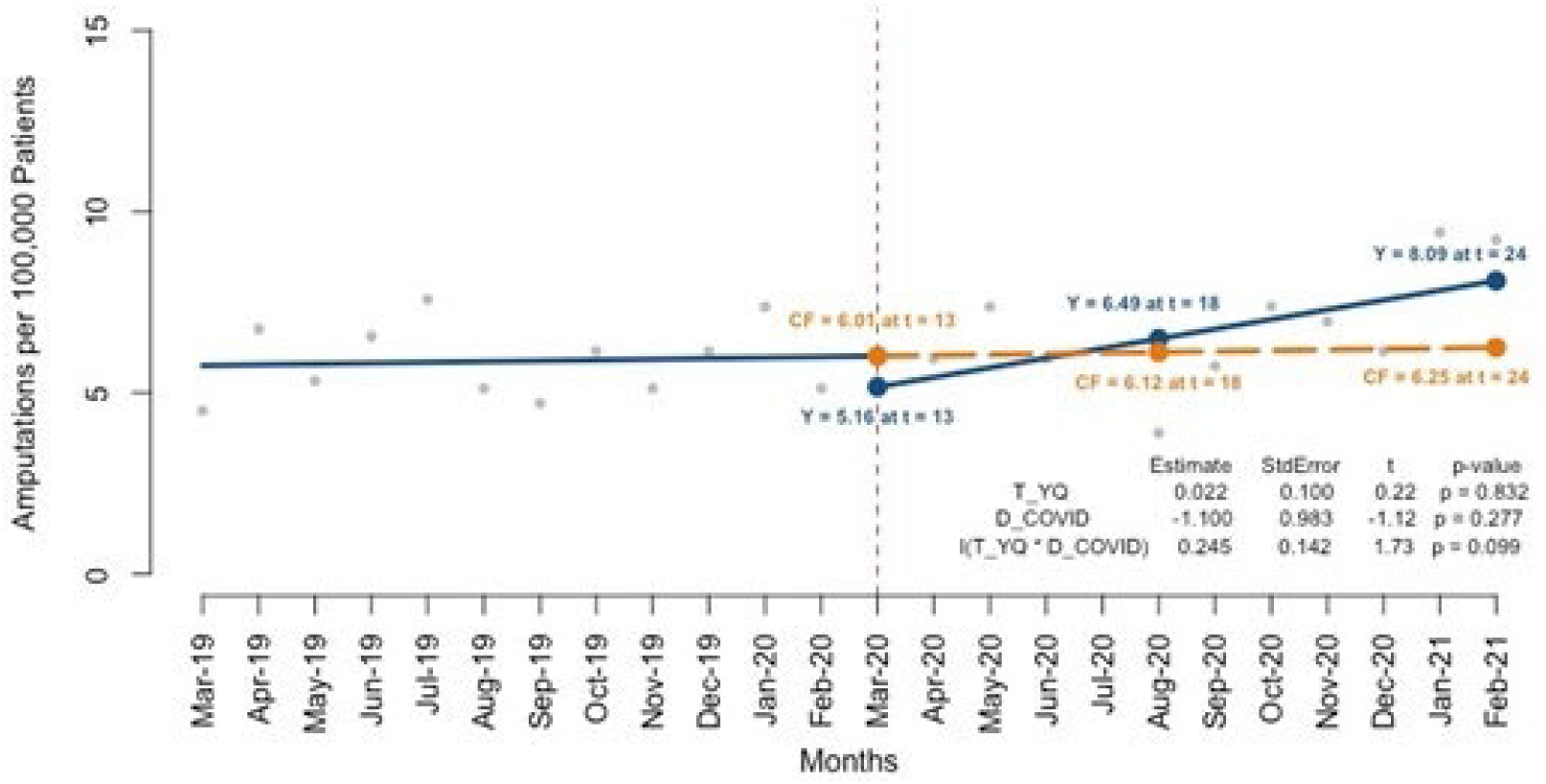
Amputations per 100,000 Type 2 Diabetes Patients Aged 65+ Relative drop = 13.9%

Level of education also influenced amputation outcomes in individuals with type 2 diabetes. Patients with a high education level showed a borderline significant increase in amputation rates post-pandemic (0.296, p=0.064) (Figure 3a1), whereas those with low education level experienced a significant increase (0.479, p=0.043) (Figure 3a2). Income disparities were evident, with patients earning less than $40,000 per year showing a significant increase in amputation rates post-COVID-19 (0.689, 0.027) (Figure 3b1). Those in the $40,000-$125,000 income bracket exhibited borderline significant increase (0.288, p=0.052) (Figure 3b2), while patients earning more than $125,000 showed no significant changes (p=0.569) (Figure 3b3).

**Figure 3:**
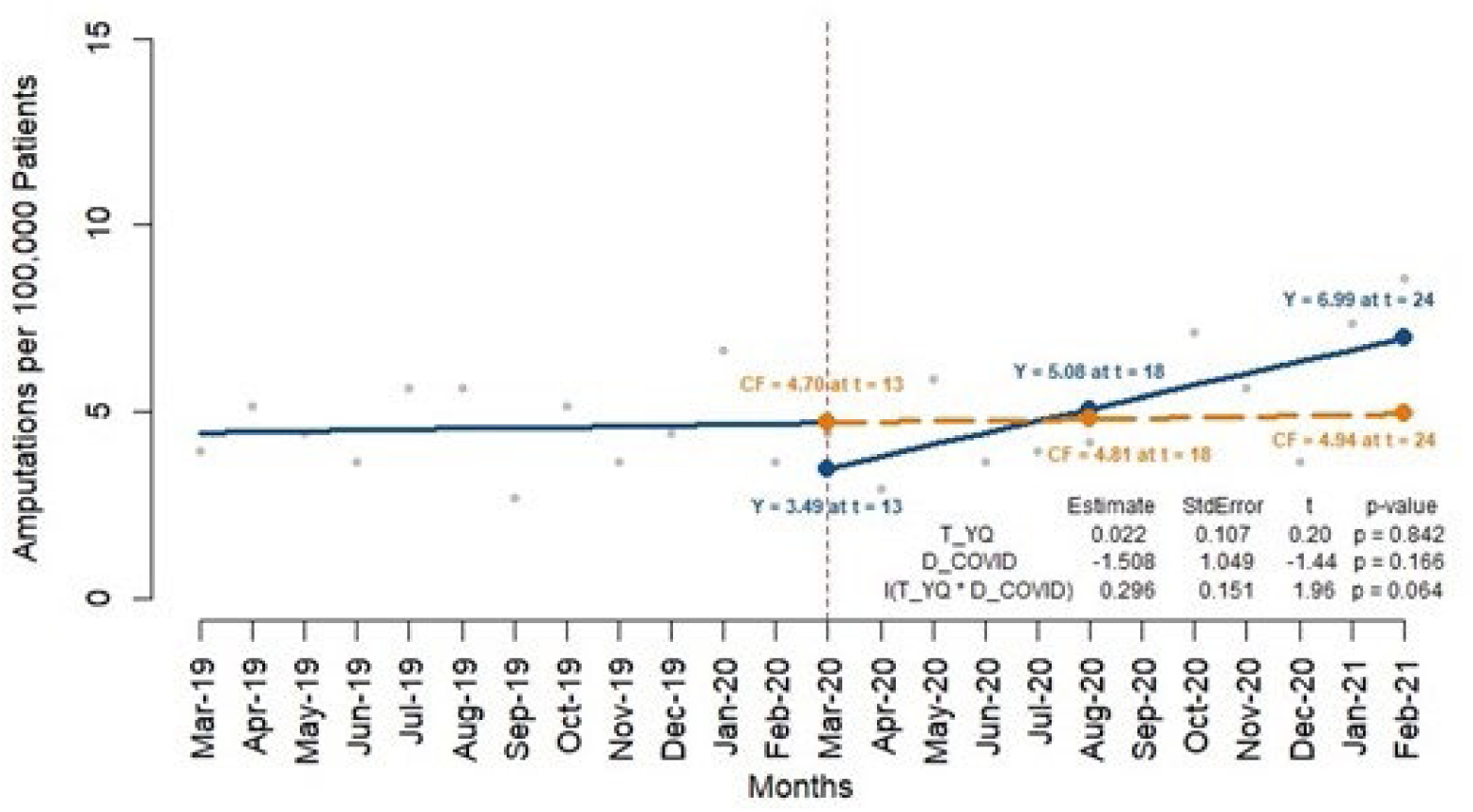
Amputations per 100,000 Type 2 Diabetes Patients by Social Determinants of Health. **Figure 3a1:** Amputations per 100,000 Type 2 Diabetes Patients with higher education Relative drop = 25.4%

**Figure 3a2:**
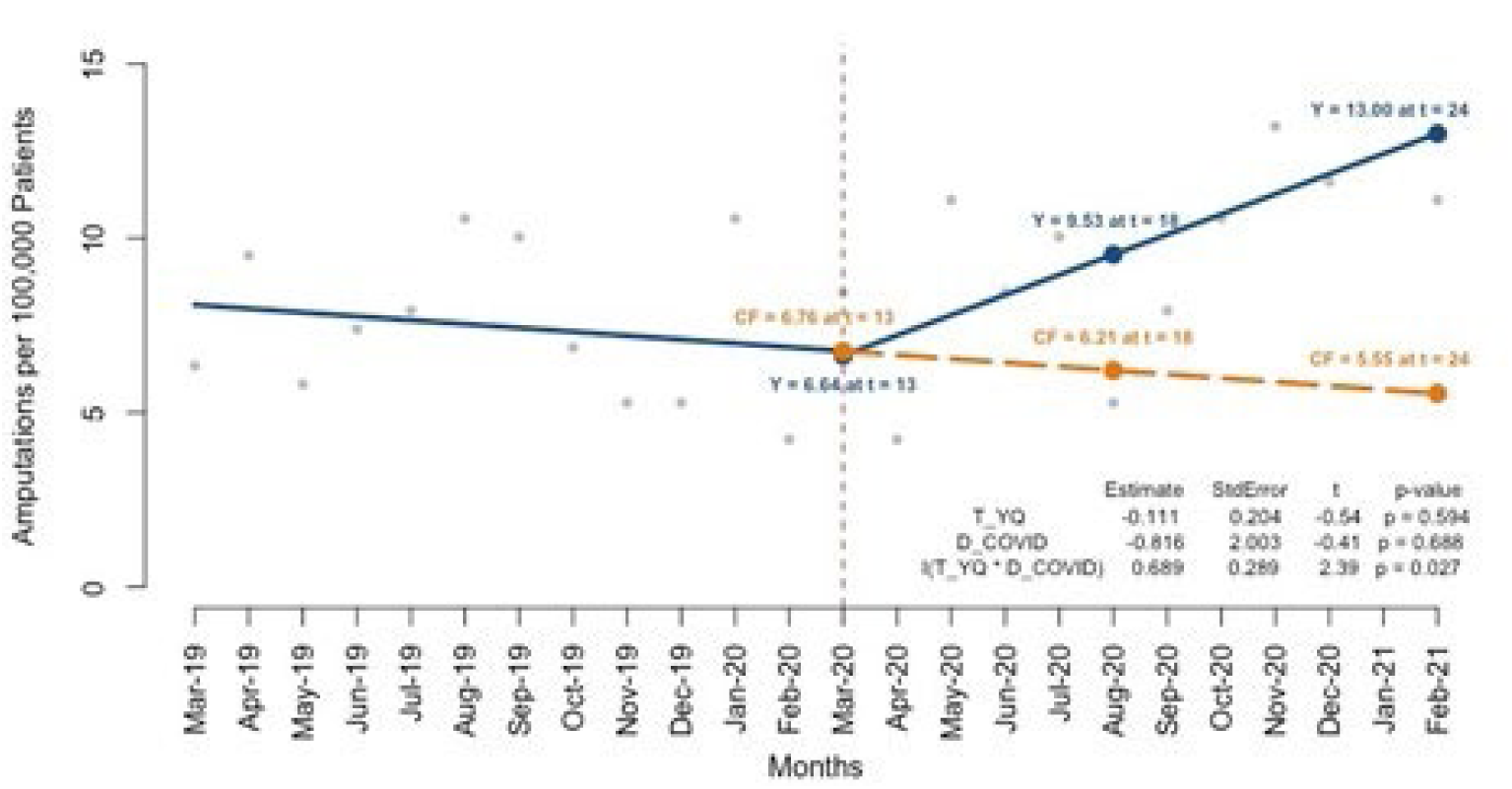
Amputations per 100,000 Type 2 Diabetes Patients with lower education Relative drop = 11.2%

**Figure 3b1:**
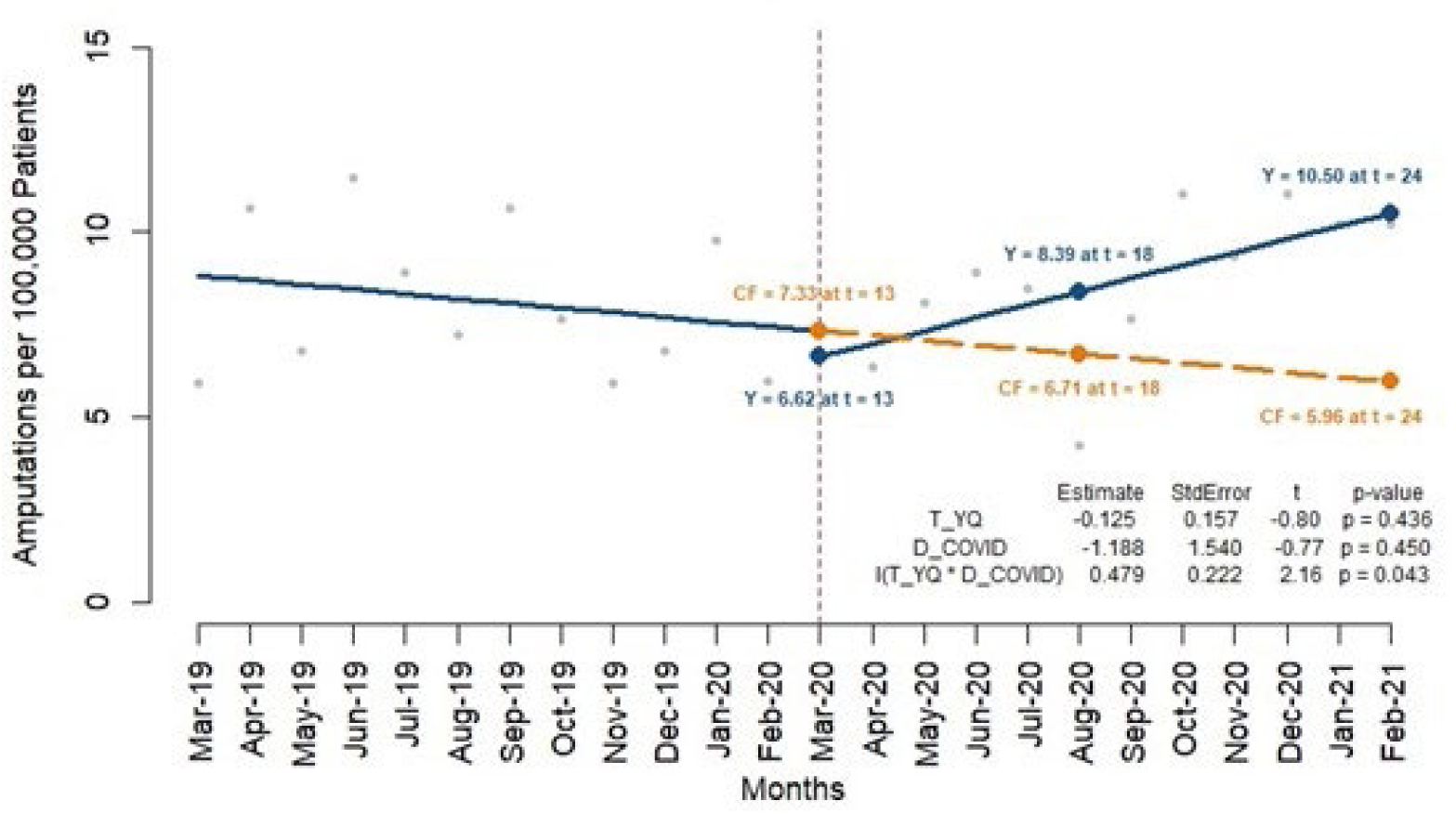
Amputations per 100,000 Type 2 Diabetes Patients with income <$40,000 Relative drop = 3.5%

**Figure 3b2:**
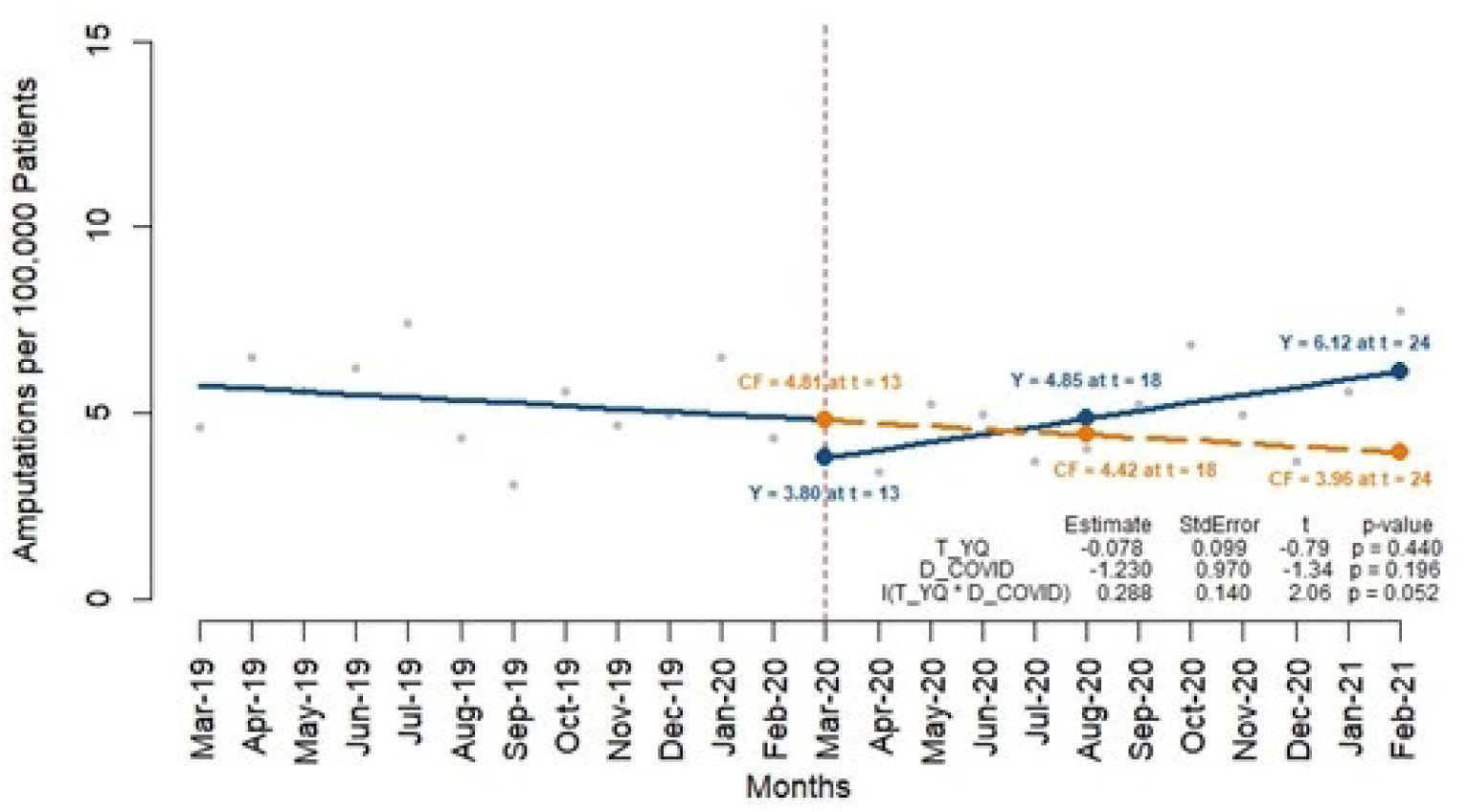
Amputations per 100,000 Type 2 Diabetes Patients with income $40,000 -$125,000 Relative drop = 22.3%

**Figure 3b3:**
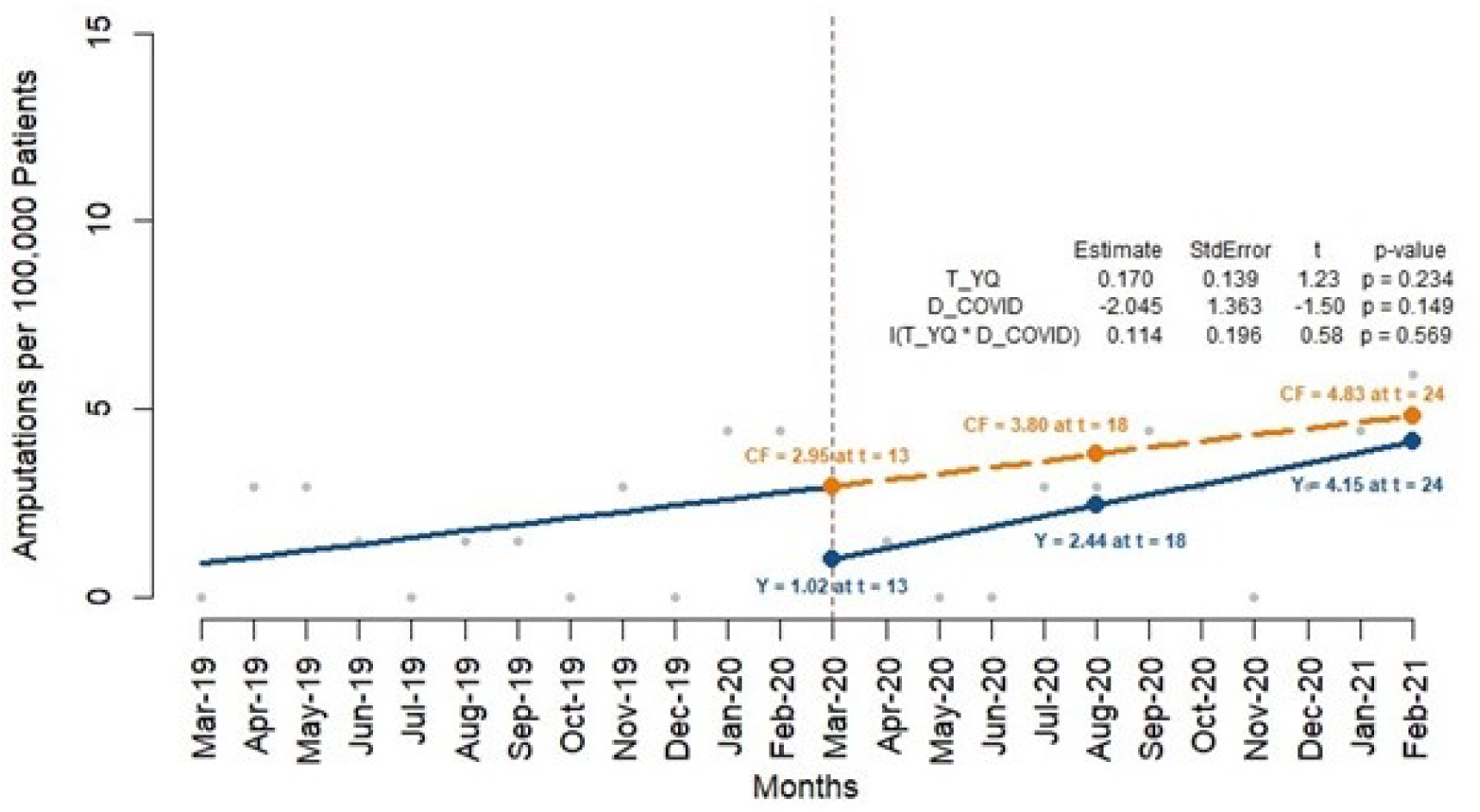
Amputations per 100,000 Type 2 Diabetes Patients with income >$125,000 Relative drop = 63.3%

**Figure 3c1:**
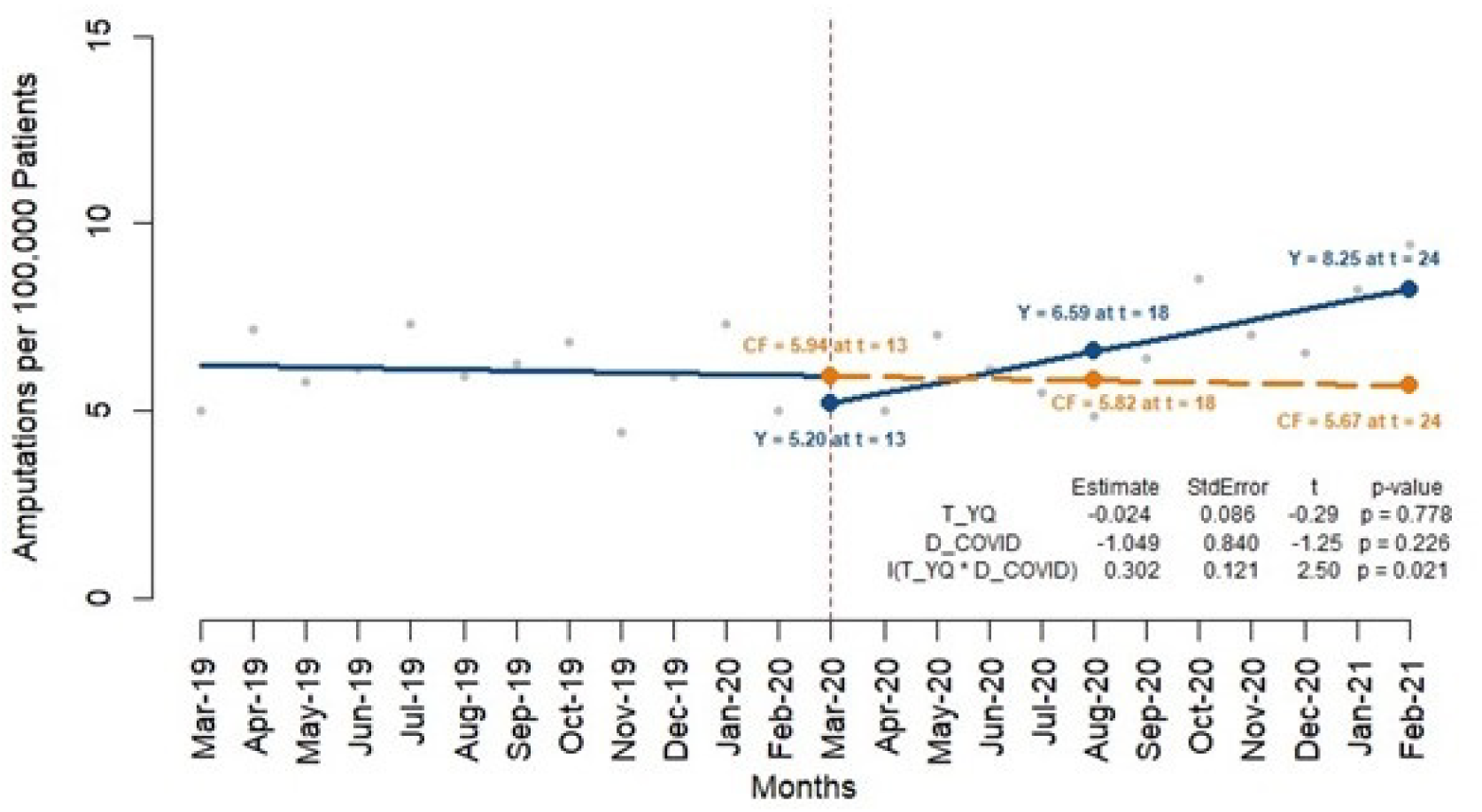
Amputations per 100,000 Type 2 Diabetes Patients in Metropolitan/Micropolitan Areas Relative drop = 12.9%

**Figure 3c2:**
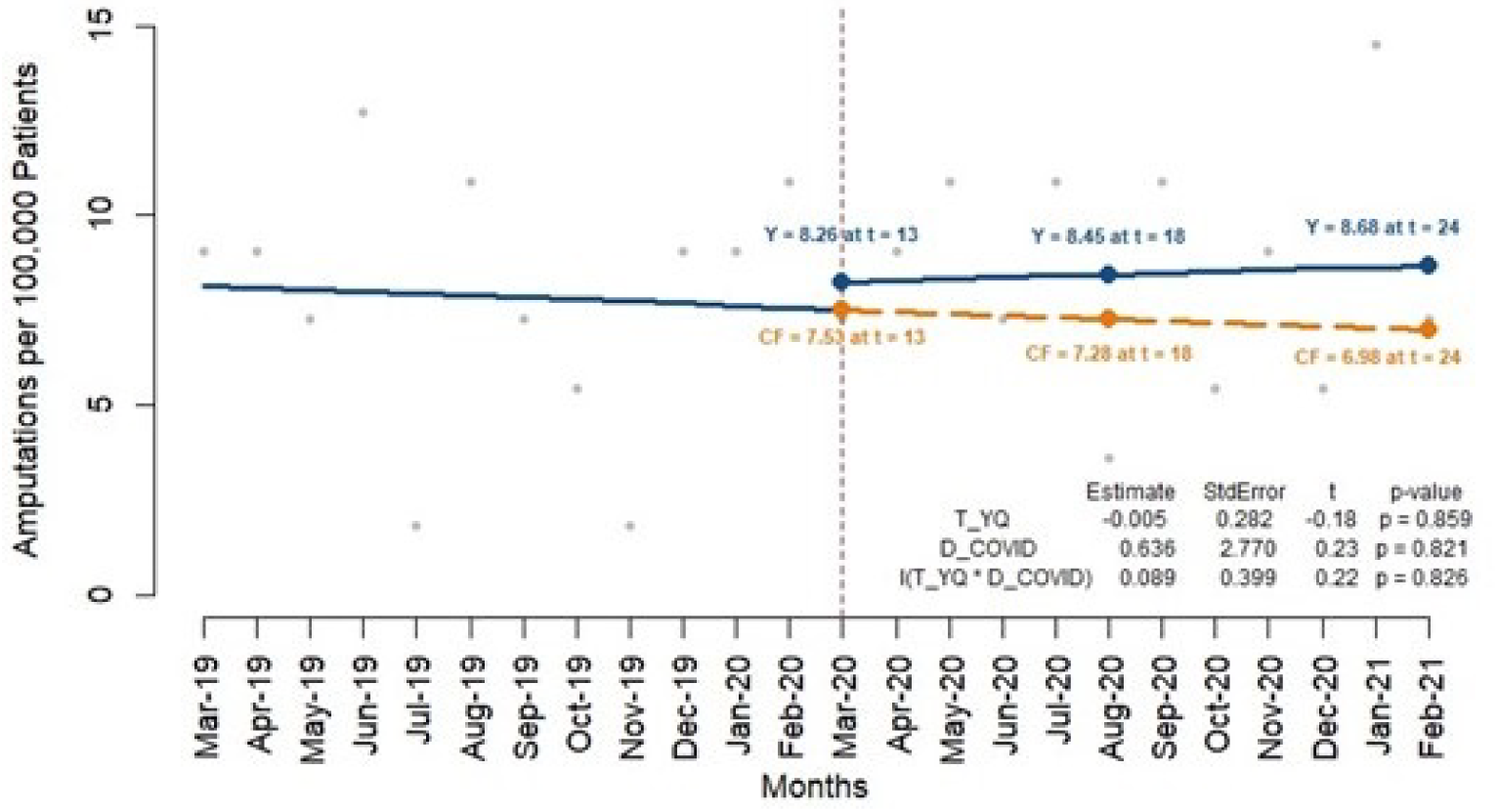
Amputations per 100,000 Type 2 Diabetes Patients in Small Town/Rural Areas Relative increase = 8.9%

Geographic locations further highlighted disparities, with patients living in metropolitan areas experiencing a significant increase in amputation rates (0.302, p=0.021) (Figure 3c1). In contrast, no significant changes were observed for patients in small towns or rural areas (p=0.826) (Figure 3c2).

White patients experienced a significant increase in amputation rates post-COVID-19 (0.343, p=0.032) (Figure 4a). In contrast, non-White patients did not exhibit any significant changes in amputation rates during the same period (p=0.277) (Figure 4b).

**Figure 4:**
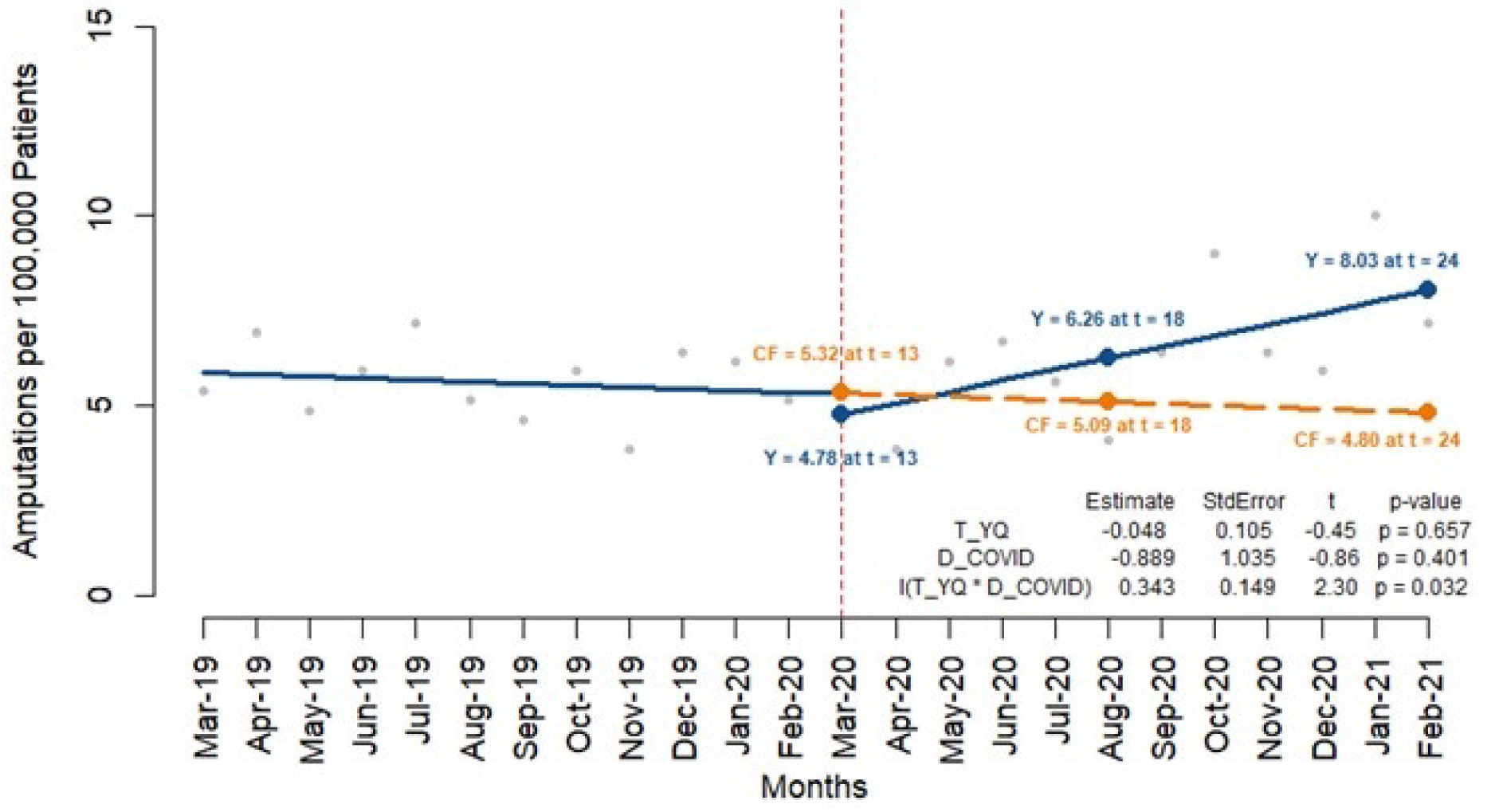
Amputations per 100,000 Type 2 Diabetes Patients by Race/Ethnicity. **Figure 4a:** Amputations per 100,000 Type 2 Diabetes Patients (non-Hispanic Whites) Relative drop = 11.0%

**Figure 4b:**
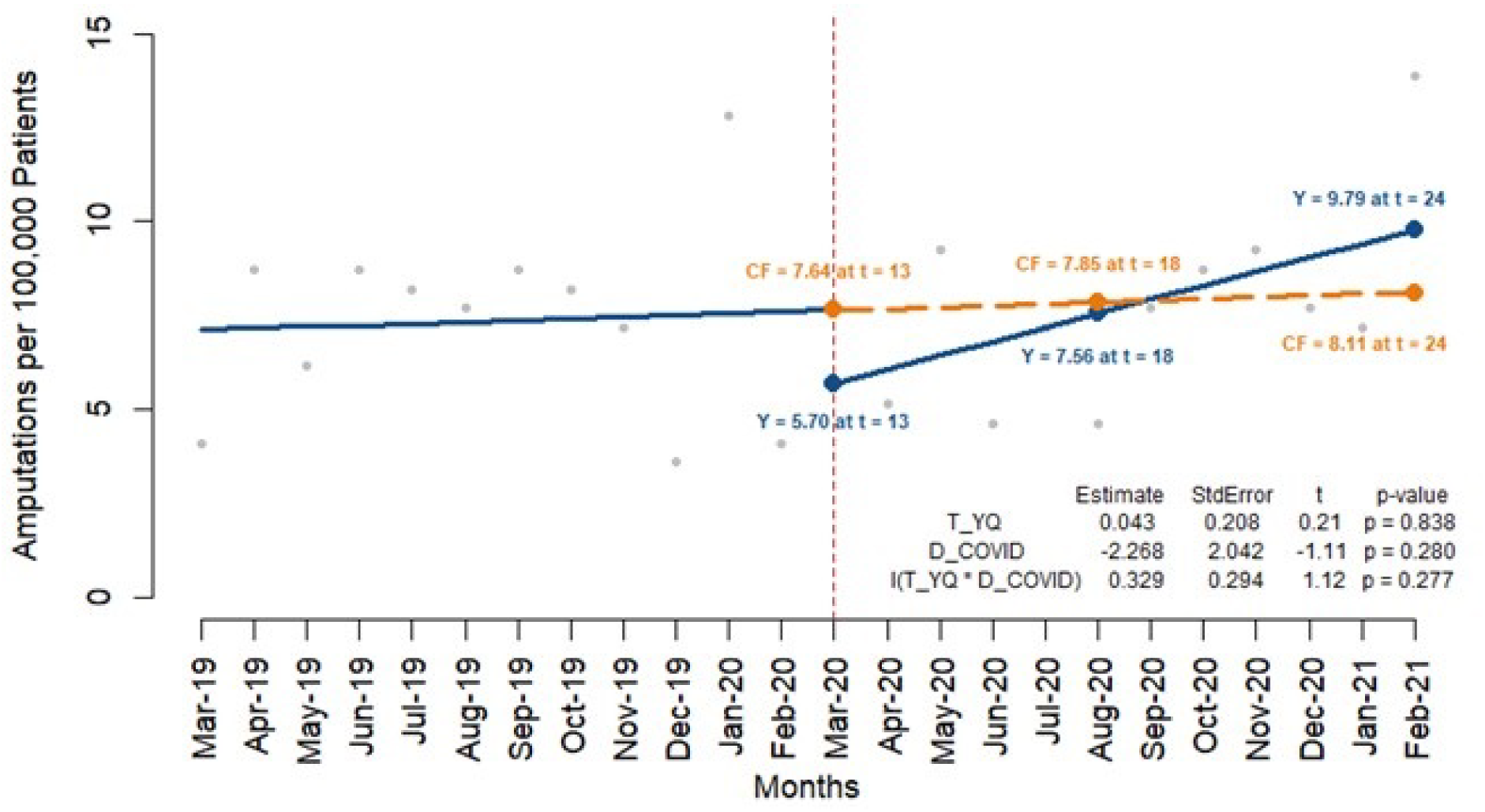
Amputations per 100,000 Type 2 Diabetes Patients (non-Whites) Relative drop = 25.0%

## DISCUSSION & CONCLUSION

The analysis of amputation rates for individuals with type 1 and type 2 diabetes revealed significant differences before and after the onset of COVID-19 pandemic. These findings highlight a notable disruption in healthcare access for those with type 2 diabetes, characterized by an initial decline in amputations followed by a compensatory increase as delayed care needs were addressed. In contrast, the amputation rates for type 1 diabetes patients showed no significant fluctuations, indicating a different pattern of healthcare utilization and outcomes during the pandemic. Only one other study examine the rate of DM related foot problems, including amputation during the COVID-19 pandemic. Their 1-year retrospective chart review showed the odds of undergoing any amputation was 10.8 times higher during the pandemic versus before the pandemic and 3.1 times higher for major amputations.^10^

The different impact of the pandemic on individuals with type 1 and type 2 diabetes may be attributed to the varying complexities in disease management and healthcare access challenges specific to each group. Those with type 1 diabetes often have more stringent and continuous monitoring requirements, which might have led to more consistent healthcare access despite the pandemic. Conversely, those with type 2 diabetes, who typically represent a larger demographic experienced greater disruption in care, leading to significant changes in amputation rates.

The impact of social determinants on amputation outcomes was also more pronounced for individuals with type 2 diabetes during the pandemic. Patients from lower-income brackets and those with lower education levels experienced significant increases in amputation rates. These disparities suggest that the pandemic exacerbated existing inequalities in healthcare access and outcomes. Even before the pandemic, studies demonstrate that those in the lowest income quartile were at 34% higher odds of undergoing an amputation compared to those in the highest income quartile.^11^ Specifically, lower-income individuals likely faced heightened financial barriers to accessing care, while those with lower education levels might have encountered greater difficulties navigating healthcare system during the pandemic crisis. Metropolitan residents, possibly affected by higher rates of COVID-19 infection and more stringent lockdown measures, also saw significant disruptions in healthcare access.

Our analysis also revealed notable patterns in the relative drops in amputation rates across different demographic and socioeconomic groups during the initial onset of the COVID-19 pandemic. For individuals with T1DM, the relative drop in amputation rates was 24.7%, with more pronounced decreases observed in older age groups (46.0% for those 65+), those with lower socioeconomic status (35.5% for income <$40k), and non-white patients (42.4%). Individuals with T2DM experienced a smaller overall relative drop of 10.9%, with the most substantial reductions seen in patients with higher incomes (63.3% for income >$125k), higher education level (25.4% for income > $125k) and non-white patients (25.4%). This pattern indicates that, individuals with type 2 diabetes in higher socioeconomic groups were more likely to defer care, possibly due to greater health awareness and risk aversion.

These findings highlight the urgent need for targeted interventions to address the social determinants of health that contribute to these disparities, especially during public health crises. Strategies to mitigate these effects could include improving access to telehealth services, ensuring the availability of affordable healthcare, and enhancing patient education and support systems to better navigate healthcare services during emergencies. Efforts to improve vascular health in those with diabetes should also focus on the subgroups at high risk for amputation and the disparities they face, and this need likely continues even in the current post-COVID years.^11^

This study has several limitations that should be considered. First, the small number and high variability of amputations, particularly among those with type 1 diabetes, may have affected our ability to detect significant changes in this group. Second, the study period may have been too short to capture the full impact of the pandemic on amputation outcomes. Amputations often result from long-term complications and are exacerbated by delayed care. Observing these amputation outcomes over a longer period might have revealed more significant changes and provided a more comprehensive understanding of the pandemic’s impact.

In conclusion, the COVID-19 pandemic significantly impacted amputation outcomes among individuals with diabetes, with those with type 2 diabetes and those from disadvantaged social groups being most affected. Addressing these disparities requires a concerted effort to improve healthcare access for vulnerable populations, particularly during times of crisis.

## Data Availability

All data produced in the present study are available upon reasonable request to the senior author, Leslie Wilson.

## Contributions from non-authors

- None

## Disclaimers

- None

## Author Contributions

Yelena Ionova: Conceptualization, Data curation, Formal analysis, Investigation, Methodology, Software, Roles/Writing – original draft

Lixian Zhong: Conceptualization, Investigation, Methodology, Writing – review & editing

Ruben Vargas: Data curation, Formal analysis, Investigation, Methodology, Software, Validation, Visualization, Writing – review & editing

Yanlei Ma: Conceptualization, Investigation, Methodology, Writing – review & editing

Leslie Wilson: Conceptualization, Data curation, Formal analysis, Funding acquisition, Methodology, Project administration, Resources, Software, Supervision, Writing – review & editing

## Previous Presentations

- None

