## Supplemental for "COVID-19 Impact on Individuals with Type 1 and Type 2 Diabetes: Comparison of Pre- and Post-COVID-19 Hospitalizations for Diabetes-Related Amputation"

### Supplemental Materials

**Supplemental Table 1: Demographic Characteristics of Study Population**

| Demographic | Type 1 Diabetes (N = 103,290) | Type 2 Diabetes (N = 711,951) |
|---|---|---|
| **Age** | | |
| 18-44 years | 14,058 (13.6%) | 26,588 (3.7%) |
| 45-64 years | 31,433 (30.4%) | 197,183 (27.7%) |
| 65+ years | 57,799 (65.0%) | 488,180 (68.6%) |
| **Gender** | | |
| Female | 57,752 (51.1%) | 365,446 (51.3%) |
| Male | 50,535 (48.9%) | 346,475 (48.7%) |
| **Race/Ethnicity** | | |
| Asian | 2,302 (2.2%) | 28,936 (4.1%) |
| Black | 17,659 (17.1%) | 106,236 (14.9%) |
| Hispanic | 11,311 (11.0%) | 88,599 (12.4%) |
| White | 56,568 (54.8%) | 389,158 (54.7%) |
| Unknown/Did not provide answer | 15,450 (15.0%) | 99,022 (13.9%) |
| **Income** | | |
| <$40,000 | 28,745 (27.8%) | 189,341 (26.6%) |
| $40,000-$74,999 | 23,413 (22.7%) | 181,410 (25.5%) |
| $75,000-$124,999 | 18,042 (17.5%) | 141,306 (19.8%) |
| $125,000-$199,999 | 6,975 (6.8%) | 48,489 (6.8%) |
| >$200,000 | 3,868 (3.7%) | 19,248 (2.7%) |
| Unknown/Did not provide answer | 22,247 (21.5%) | 132,157 (18.6%) |
| **Education** | | |
| Less than 12$^{th}$ Grade | 391 (0.4%) | 3,362 (0.5%) |
| High School | 34,108 (33.0%) | 232,002 (32.6%) |
| Some college | 46,774 (45.3%) | 336,491 (47.3%) |
| Bachelor's or Higher | 11,263 (10.9%) | 70,767 (9.9%) |
| Unknown/Did not provide answer | 10,754 (10.4%) | 69,329 (9.7%) |
| **Rurality** | | |
| Metropolitan | 84,831 (82.1%) | 587,807 (82.6%) |
| Micropolitan | 10,069 (9.7%) | 67, 977 (9.5%) |
| Small town | 5,517 (5.3%) | 36,509 (5.1%) |
| Rural area | 2,713 (2.6%) | 18,608 (2.6%) |
| Unknown/Did not provide answer | 160 (0.2%) | 1,050 (0.1%) |

**Supplement Figure 1: Flow chart of the patient selection (T1DM and T2DM)**

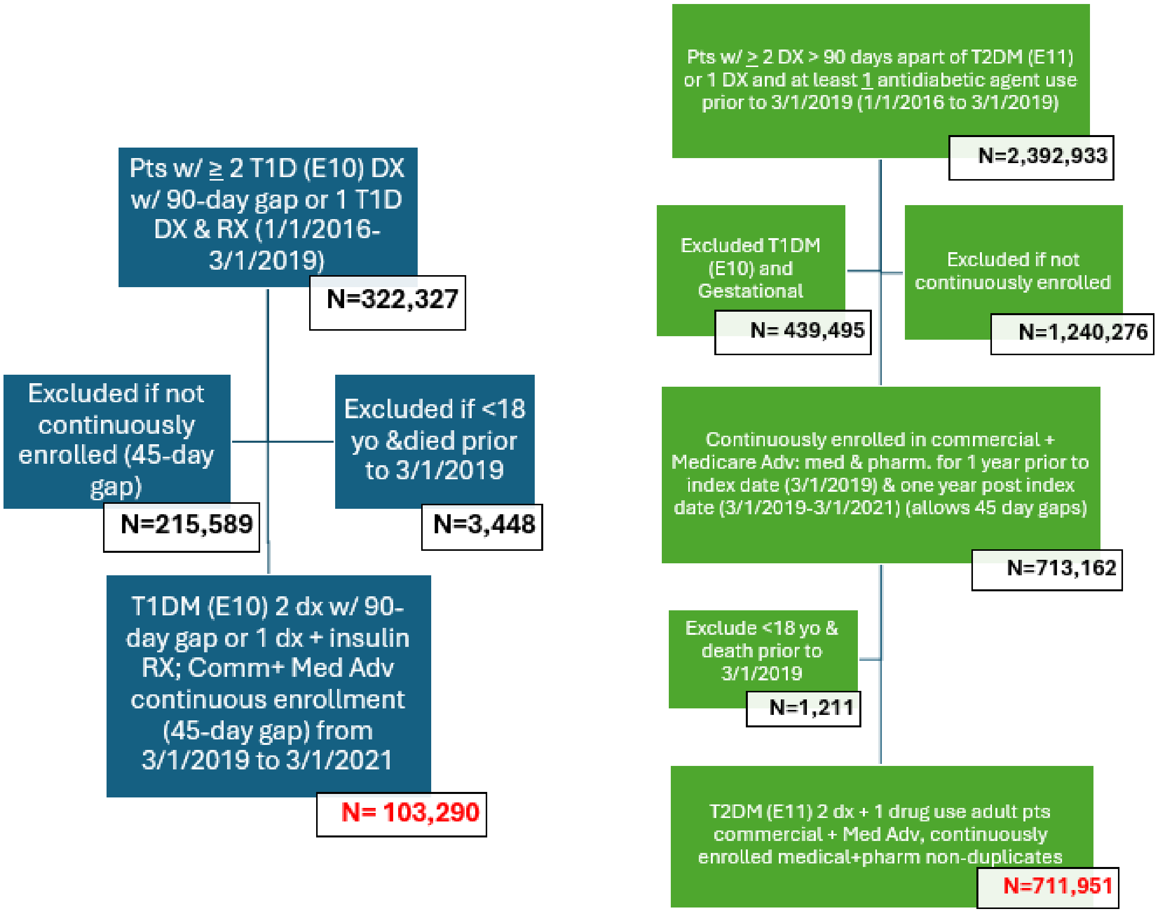

**Supplemental Figure 2a: Amputations per 100,000 Type 1 Diabetes Patients with higher education**

Relative drop = 34.2%

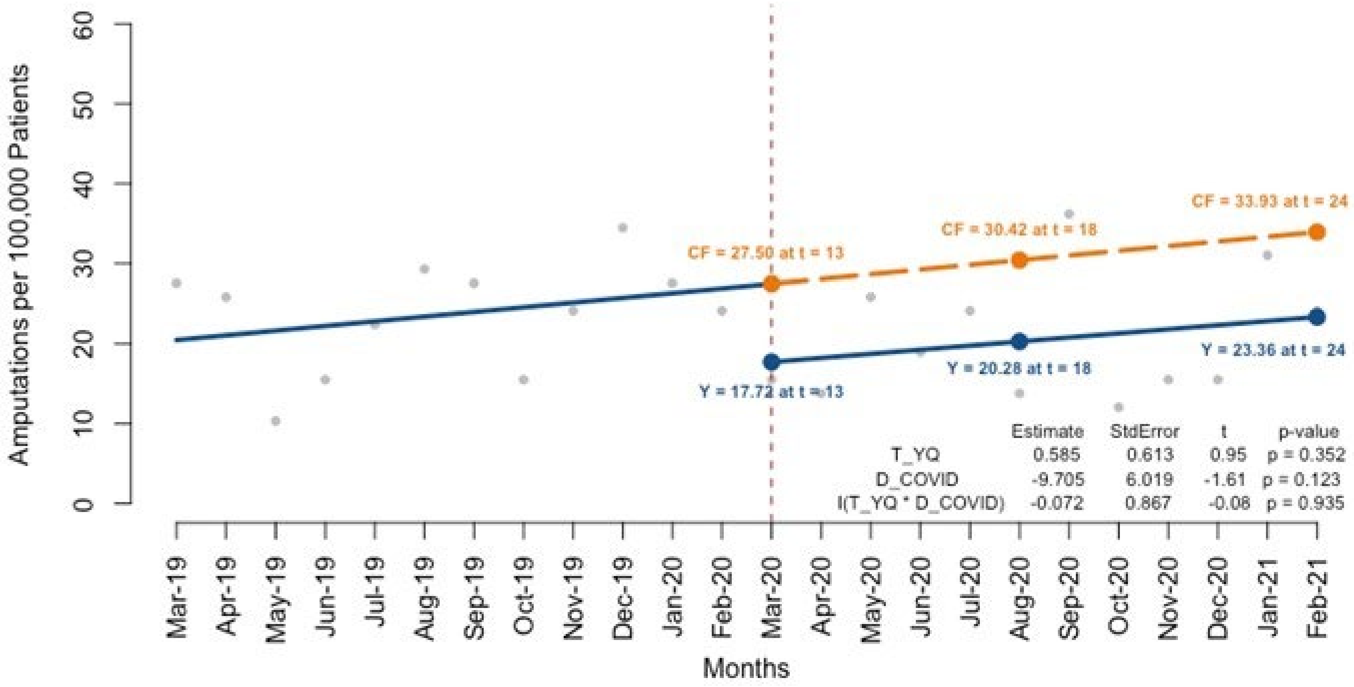


**Supplemental Figure 2b: Amputations per 100,000 Type 1 Diabetes Patients with lower education**

Relative drop = 26.1%

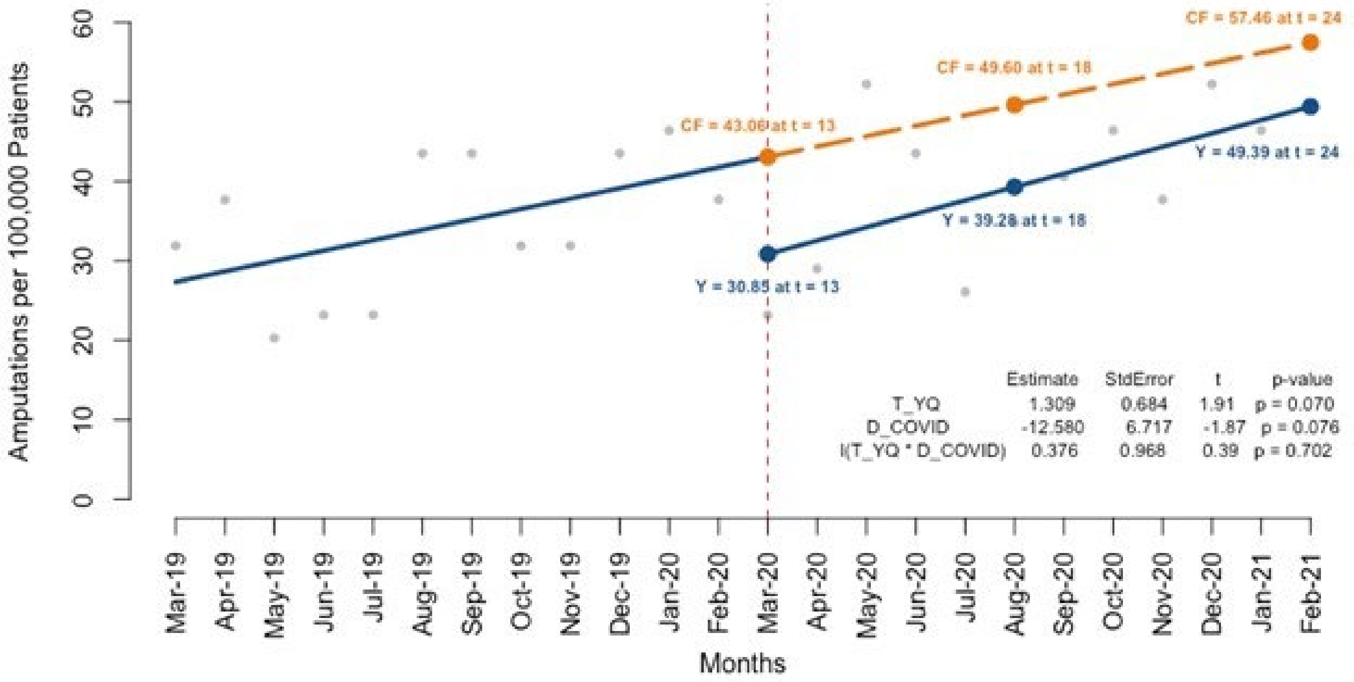

**Supplemental Figure 3a: Amputations per 100,000 Type 1 Diabetes Patients (non-Hispanic Whites)**

Relative drop = 23.1%

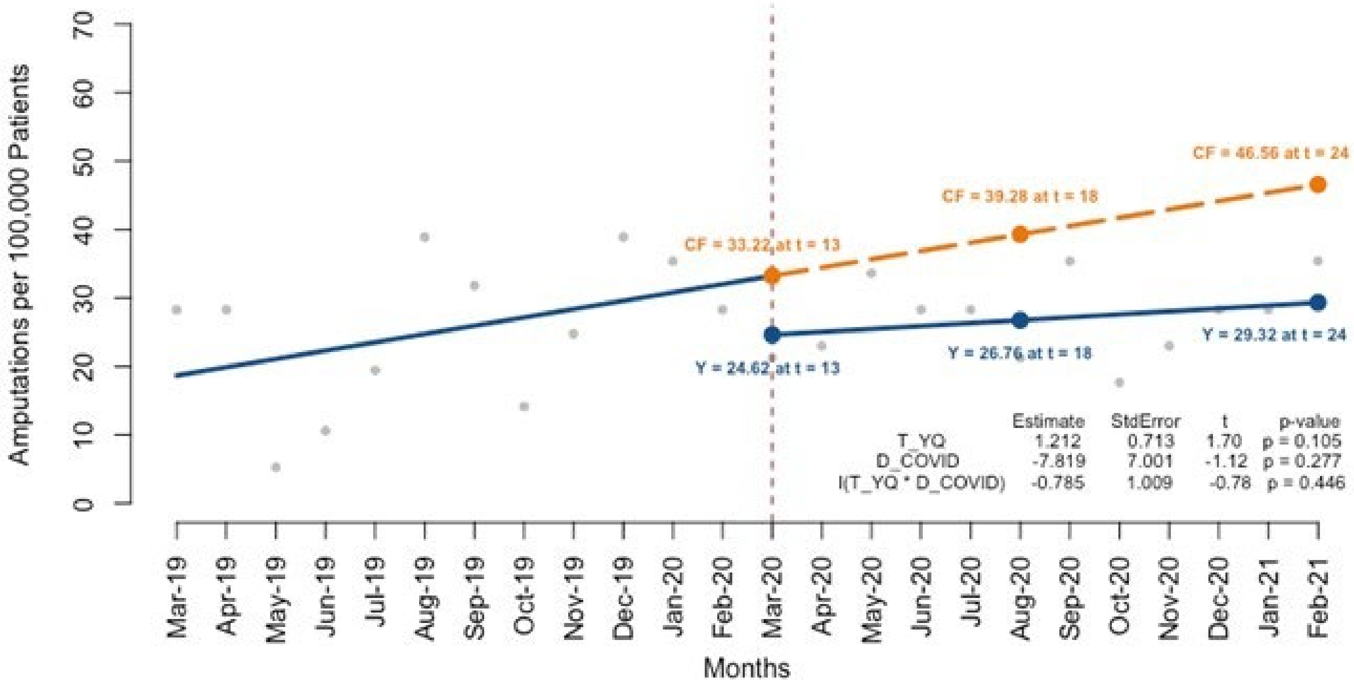


**Supplemental Figure 3b: Amputations per 100,000 Type 1 Diabetes Patients (non-Whites)**

Relative drop = 42.4%

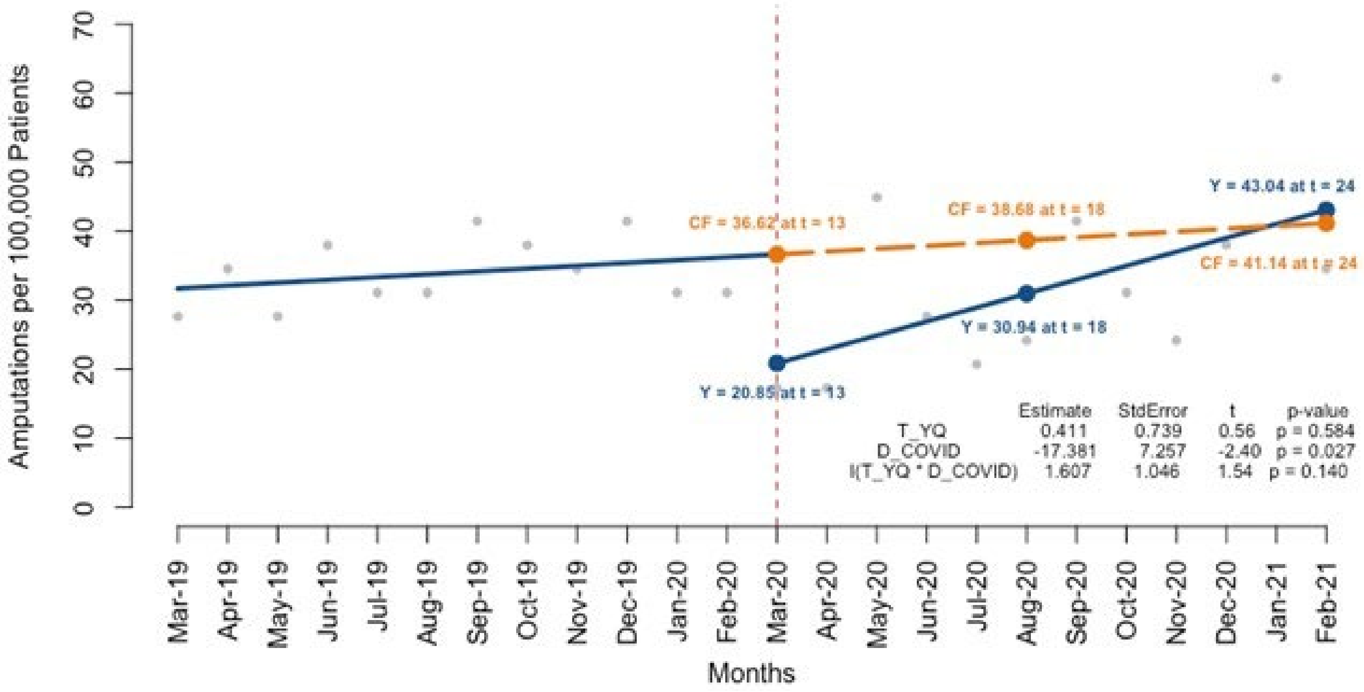

**Supplemental Figure 4a: Amputations per 100,000 Type 1 Diabetes Patients Aged 18-44**

Relative increase = 2.4%

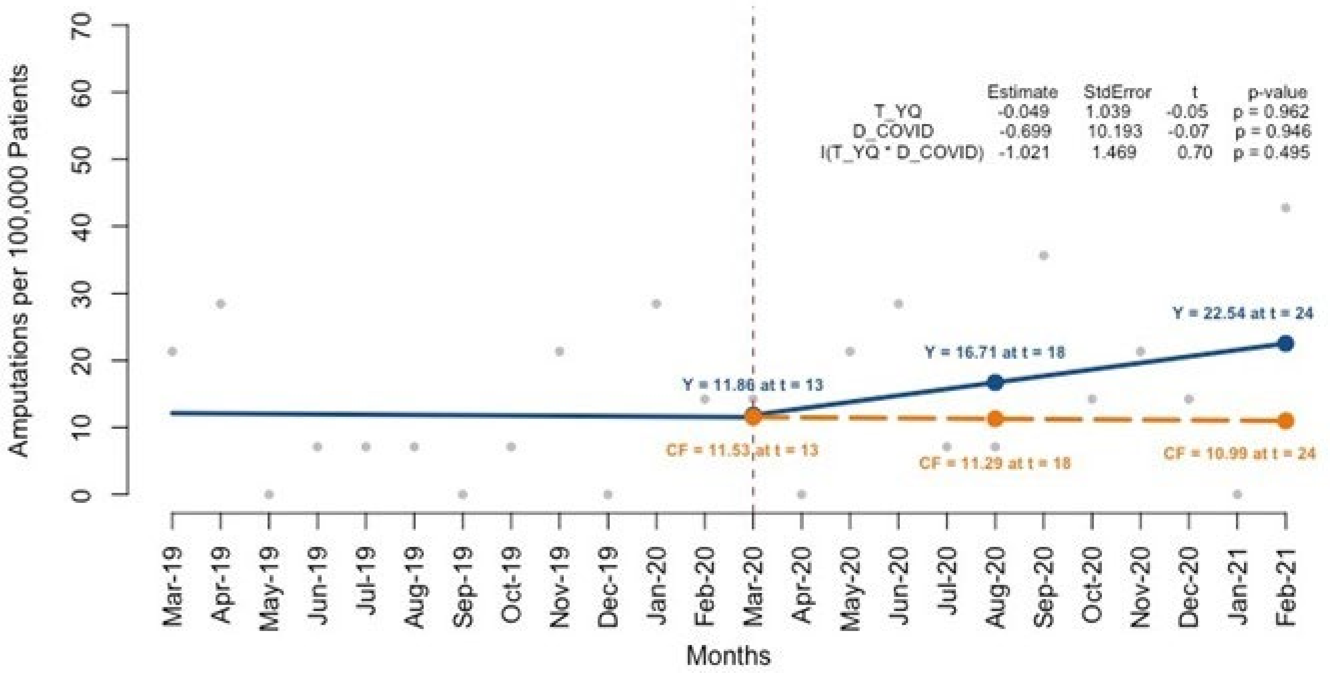


**Supplemental Figure 4b: Amputations per 100,000 Type 1 Diabetes Patients Aged 45-64**

Relative drop = 2.4%

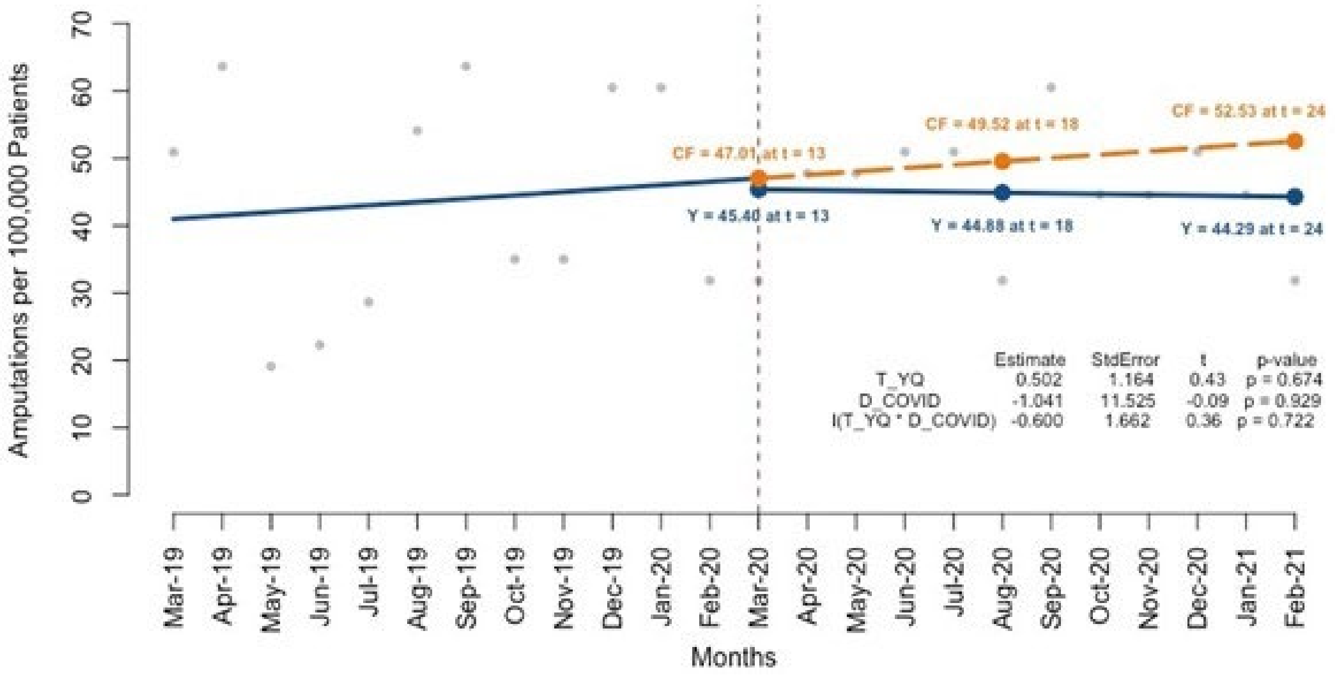

**Supplemental Figure 4c: Amputations per 100,000 Type 1 Diabetes Patients Aged 65+**

Relative drop = 46.0%

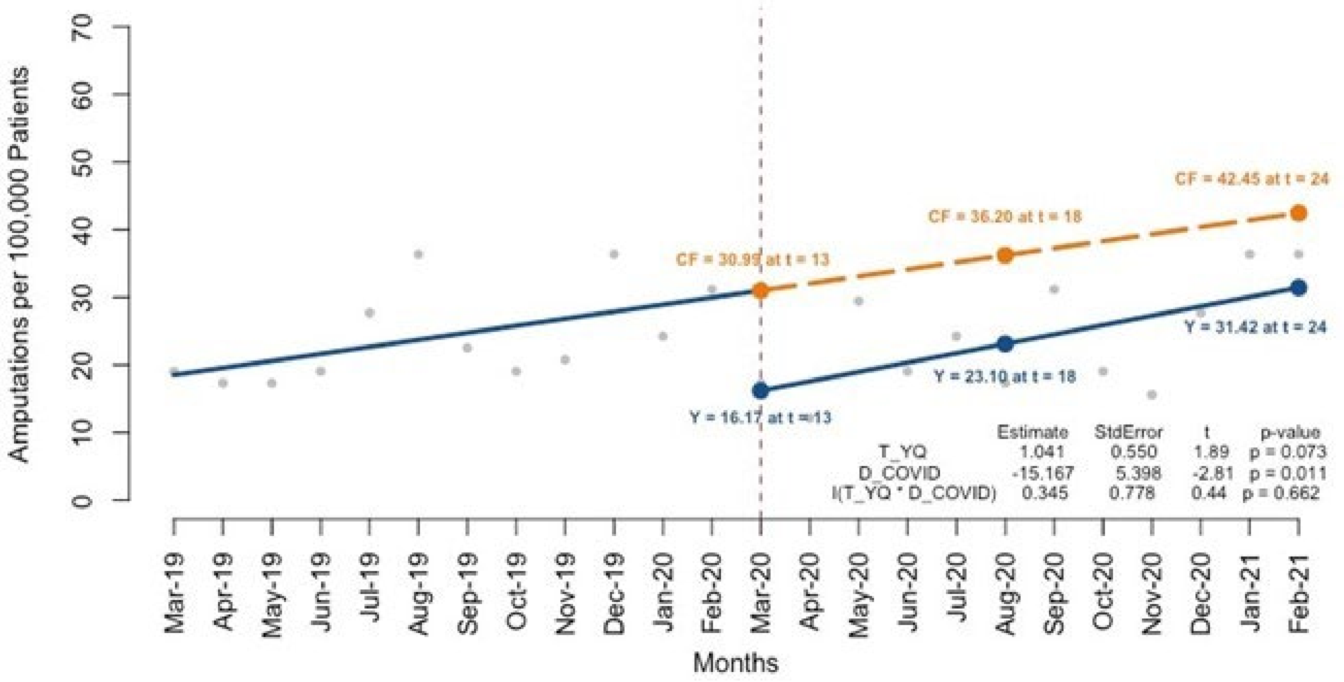


**Figure 5a: Amputations per 100,000 Type 1 Diabetes Patients with income <$40,000**

Relative drop = 35.5%

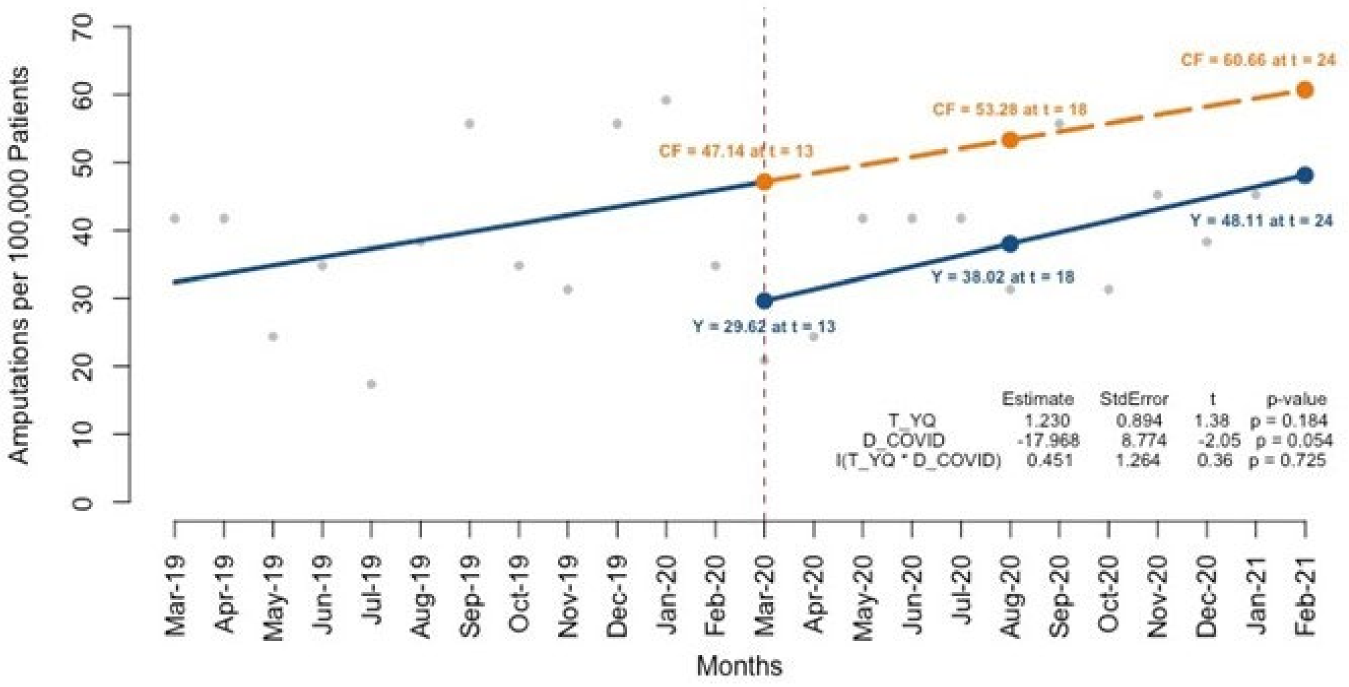

**Figure 5b: Amputations per 100,000 Type 1 Diabetes Patients with income $40,000 - $125,000**

Relative drop = 42.0%

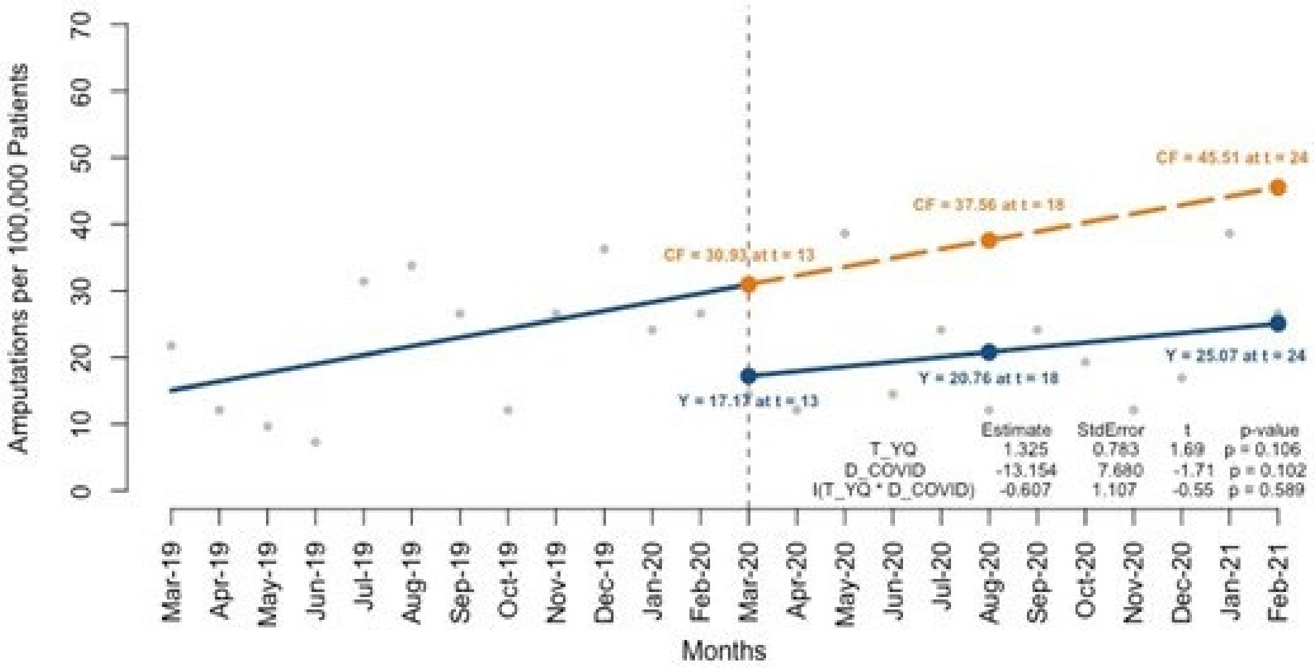


**Supplemental Figure 5c: Amputations per 100,000 Type 1 Diabetes Patients with income >$125,000**

Relative drop = 26.1%

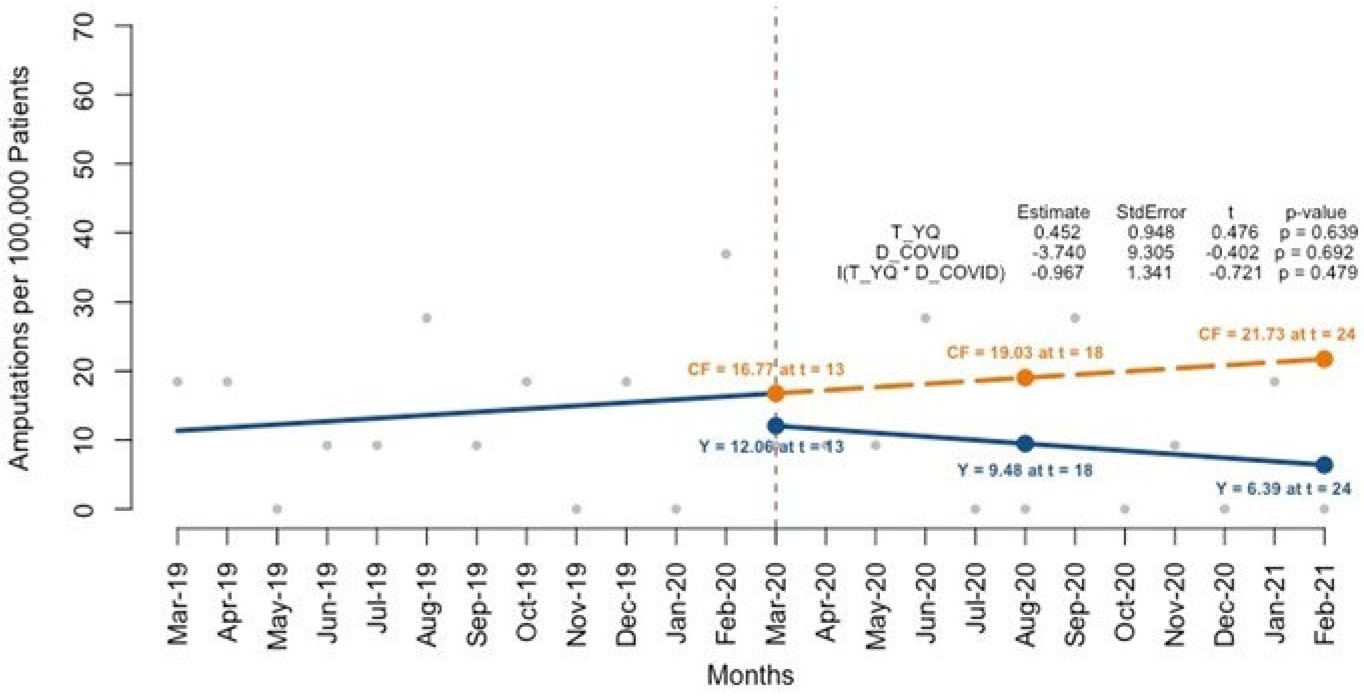

**Supplemental Figure 6a: Amputations per 100,000 Type 2 Diabetes Patients in Metropolitan/Micropolitan Areas**

Relative drop = 30.2%

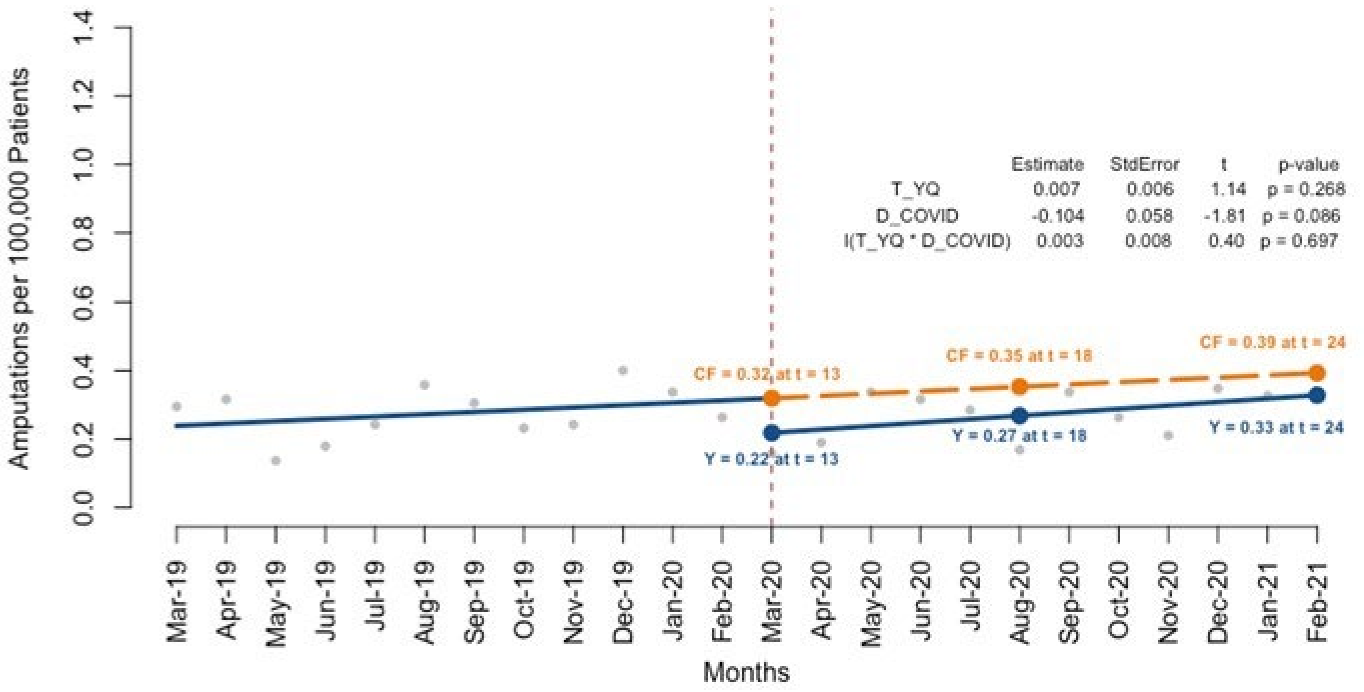


**Supplemental Figure 6b: Amputations per 100,000 Type 2 Diabetes Patients in Small Town/Rural Areas**

Relative increase = 17.5%

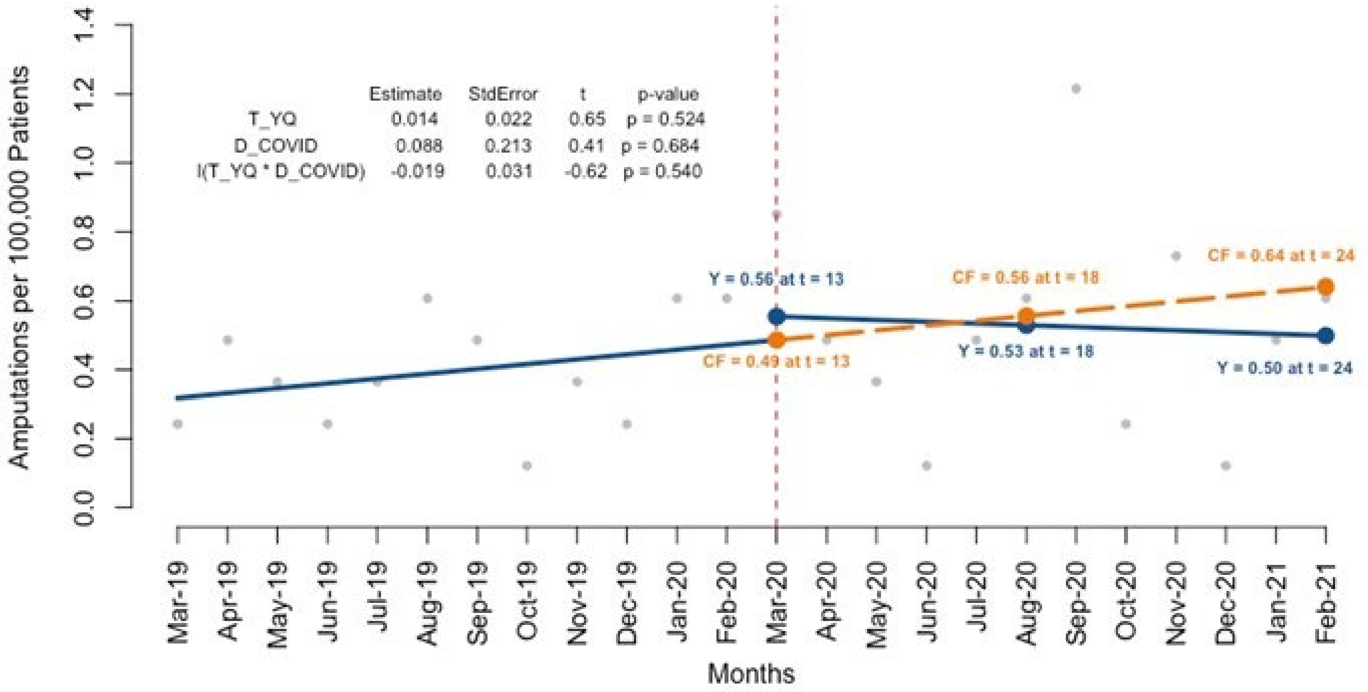
